# Cross-sectional study of plasma phosphorylated Tau 217 in persons without dementia

**DOI:** 10.1101/2024.05.17.24307528

**Authors:** Toni T. Saari, Teemu Palviainen, Mikko Hiltunen, Sanna-Kaisa Herukka, Tarja Kokkola, Sari Kärkkäinen, Mia Urjansson, Aino Aaltonen, Aarno Palotie, Heiko Runz, Jaakko Kaprio, Valtteri Julkunen, Eero Vuoksimaa, FinnGen

**Affiliations:** Institute for Molecular Medicine Finland (FIMM), Helsinki Institute of Life Science, University of Helsinki, Finland; Institute of Biomedicine, University of Eastern Finland, Kuopio, Finland; Institute of Clinical Medicine/Neurology, University of Eastern Finland, Kuopio, Finland; Department of Neurology, Neurocenter, Kuopio University Hospital, Kuopio, Finland; Analytic and Translational Genetics Unit, Department of Medicine, Department of Neurology and Department of Psychiatry Massachusetts General Hospital, Boston, MA, USA; The Stanley Center for Psychiatric Research and Program in Medical and Population Genetics, The Broad Institute of MIT and Harvard, Cambridge, MA, USA; European Molecular Biological Laboratories (EMBL), Heidelberg, Germany

**Keywords:** Alzheimer’s disease, Apolipoprotein E, genome-wide association study, plasma biomarkers, twins

## Abstract

**Importance:** Blood levels of phosphorylated tau 217 (p-tau217) as measured by commercially available immunoassays are an emerging tool to assess Alzheimer’s disease (AD) pathology. Little is known about the performance of p-tau217 and contributors to its variability in aged individuals without AD.

**Objective:** To determine plasma p-tau217, its associations with age, sex, education and genetic risk, to estimate heritability, and to conduct a genome-wide association study (GWAS).

**Design:** Cross-sectional observational clinical biobank recall study called TWINGEN.

**Setting:** Recall of participants in the older Finnish Twin Cohort study without AD and other diagnoses affecting cognition based on health registry data. Sample collection in March 2023– December 2023 in six study sites across Finland. Biomarkers were determined in January 2024 and data were analyzed in March–April 2024.

**Participants:** A population-based sample of 65-85 years old twins.

**Exposures:** Age, sex, education, *Apolipoprotein E* (*APOE*) genotype and polygenic risk score of AD (ADPRS). Monozygosity (MZ) versus dizygosity (DZ) in heritability estimation.

**Main outcomes and measures:** Plasma p-tau217 (ALZpath pTau217 assay): continuous and >0.42 pg/mL cut-off. Heritability. Single nucleotide polymorphisms (SNP’s) in the GWAS.

**Results:** The study included 697 participants (mean [SD] age, 76.2 [4.6] years); 398 [57%] women and 299 [43%] men; 240 MZ [81 full pairs], 450 DZ [73 full pairs]; and 203 [29%] *APOE* ε4-allele carriers. P-tau217 was higher as a function of age (means [SDs] from 0.32 [0.20] pg/mL in 65-69 year-olds to 0.53 [0.36] pg/mL in 80-85 year-olds). Twin-based heritability was 0.56 (95%CI 0.36-0.79). *APOE* ε4-allele carriers (mean [SD]= 0.58 [0.35] pg/mL) had higher p-tau217 than non-carriers (mean [SD]= 0.39 [0.27] pg/ml, *P* <.001). Abnormal p-tau217 levels of >0.42 pg/mL were evident in 39% and predicted by higher age (OR=1.15 [95%CI, 1.10-1.20]) and having *APOE* ε4-allele (OR=4.53 [95%CI, 3.10-6.62]). Sex, education and ADPRS were not related to p-tau217 levels or abnormality. GWAS indicated 45 SNPs associated with p-tau217 plasma levels (*P*<5x10^−08^) centered around the *APOE* locus.

**Conclusions and relevance:** Our results support the use of plasma p-tau217 as a biomarker in detecting preclinical or prodromal AD and in genome-wide association studies of biologically defined AD.

**Key points:** *Question:* To what extent are individual differences and abnormality in plasma p-tau217 associated with age, sex, and genetic factors in a population-based sample without a diagnosis of Alzheimer’s disease (AD)?

*Findings:* In this cross-sectional observational study of 697 twins (65-85-year-olds), older age and carrying an *Apolipoprotein E* (*APOE*) ε4-allele were associated with higher plasma p-tau217; abnormal levels were evident in 39% of participants. The twin-based heritability estimate was 56% and genome-wide association study implicated genes around *APOE* region.

*Meaning:* Results support using blood-based plasma p-tau217 biomarker for preclinical or prodromal AD and explain parts of its variability in aging population.

---

Advances in blood-based plasma biomarkers of Alzheimer’s disease (AD) hold a promise for cost-effective early risk evaluation and diagnosis. Recently, phosphorylated tau217 (p-tau217) has been reported to have comparable accuracy to cerebrospinal fluid biomarkers for determination of abnormal PET amyloid. ^1^ However, there is less information about plasma p-tau217 in population-based samples^2^ and the role of genetic effects in accounting for interindividual differences of the plasma protein among persons without dementia is not known.

*Apolipoprotein E* (*APOE*) gene – with ε4 as a risk allele – and polygenic risk score of AD (ADPRS) based on genome wide association study explains only a fraction of the genetic variance of clinically defined AD. ^3^ Twin studies can estimate the relative contribution of genetic and environmental effects by comparing the similarity between monozygotic (MZ, genetically identical at the sequence level) and dizygotic (DZ, genetically full siblings) twin pairs. Only one twin study has investigated the heritability of AD plasma biomarkers and found that about half of the variance in beta-amyloid (Aβ)42 and Aβ40 was explained by additive genetic effects whereas Aβ42/40 had zero heritability; also plasma total tau had 50% heritability.^4^ However, twin studies have not investigated the heritability of plasma p-tau217 despite its promise as an accurate cost-effective biomarker of both amyloid and tau positivity.^1^

The large case-control genome-wide association studies (GWAS) of AD have indicated over 80 loci.^5^ However, large samples have been reached by relaxing the criteria of clinical diagnosis and using proxy cases with familial history of dementia; thus resulting in more general dementia findings than AD specific hits.^6^ There is a paradigm shift from studying the genetics of AD based on clinical symptom diagnostics to an increased use of the biological classification of amyloid and tau pathology, correspondingly requiring new GWAS studies.^7^ Compared to clinical or proxy cases, biomarkers are more objective and reproducible measures that capture the biological process of AD. They have the added benefit of improved statistical power using continuous measures with less measurement error compared to binary clinical diagnoses.^5^ GWAS’s of AD plasma biomarkers have identified *APOE*, beta-secretase 1 (*BACE1*), presenilin 2 (*PSEN2*) and amyloid-beta precursor protein (*APP*) for plasma amyloid beta^8^ and *microtubule associated protein tau* (*MAPT*) gene for plasma total tau.^9^ Earlier GWAS studies on plasma p-tau using p-tau181 in less than 2000 individuals have implicated only *APOE* locus^10,11^ and there are no reported GWAS data of p-tau217.

Here, we conducted a population-based twin study of plasma p-tau217 to i) investigate the associations with age, sex, and education, ii) estimate the relative importance of genetic and environmental effects, iii) investigate its associations with *APOE* and ADPRS, iv) study the prevalence and predictors of abnormal levels indicative of AD pathology, and v) conduct genome-wide association study of plasma p-tau217.

## Methods

### Participants and measures

We identified twins born in 1938-1957 and included in the biobank of the Finnish Institute for Health and Welfare (THL) from the population-based older Finnish Twin Cohort (FTC) study. ^12^ After excluding those individuals with AD, other neurodegenerative disease or other cognition-affecting disease based on medical records from all hospitals in Finland, 2718 individuals (66-85 years old) were invited to participate in the TWINGEN study (see Supplement 1 for details).^13^ A total of 830 participants (31%) returned signed informed consent for participation in TWINGEN and they were contacted by phone to verify the suitability and availability for an in-person visit to one of the six study sites across Finland. Finally, 697 individuals (84% of those who returned signed consent) had an in-person study visit in March 2023 – November 2023 yielding a participation rate of 26% (697/2718).

Education was based on self-report by questionnaire. Cognitive status was based on validated telephone assessment of dementia (TELE) instrument.^14^ Genome-wide genotyping was used to define *Apolipoprotein E* (*APOE*) genotype, ^15^ and to calculate ADPRS.^16^ See eMethods in Supplement 1 for details. We chose the ADPRS balancing the proportion of variance explained, total sample size and the proportion of clinically confirmed cases and controls.^6^ To date there is no AD PRS based on biologically defined cases.

After ethical approval, all participants gave written informed consent (see eMethods for details in Supplement 1). We followed Strengthening the Reporting of Observational Studies in Epidemiology (STROBE) and STrengthening the REporting of Genetic Association Studies (STREGA) guidelines.

### Blood sample and biomarker determination

Non-fasting blood samples were drawn at one of the six study sites mostly between 9:00 and 15:00. Plasma was collected to vacutainer 10ml K2EDTA tube and then centrifuged at 1500g for ten minutes. After centrifugation, plasma was apportioned into 0.5ml aliquots and first stored at -20°C and then moved to -80°C. These aliquots were sent to the Biomarker Laboratory of the University of Eastern Finland for the biomarker analyses. P-tau217 was quantified in January 2024 using ALZpath Simoa pTau-217 v2 Assay Kit (Quanterix, Ref# 104371).^1^ Prior to analyses, EDTA plasma samples were thawed, mixed and centrifuged (10,000xg, 5 min, +20 °C).

### Genotyping

DNA was extracted from blood/saliva samples. Chip genotyping (genotype calling algorithms in parentheses) was performed using Illumina Human610-Quad v1.0 B and Human670-QuadCustom v1.0 A (Illuminus), Illumina HumanCoreExome 12 v1.0 A, 12 v1.1 A, 24 v1.0 A, 24 v1.1 A, 24 v1.2 A (GenCall), or Affymetrix FinnGen Axiom arrays (AxiomGT1). Genotype quality control was done in three batches.^17^ Prephasing was performed with Eagle v2.3^18^ and imputation with Minimac3 v2.0.1 using the University of Michigan Imputation Server.^19^ Genotypes were imputed to Haplotype Reference Consortium release 1.1 reference panel.^20^

### Statistical analyses

All analyses were done with R statistical software (version 4.3.2) in March-April 2024. Linear mixed models (*lme4* package)^21^ were used to investigate the associations of age, sex, education and *APOE* (ε4 carriers vs. non-carriers and sub-groups in post-hoc analyses) or ADPRS with p-tau217. We used logistic regression model (*survey* package)^22^ by including all these factors in predicting AD pathology based on previously published >0.42pg/ml cut-off for beta-amyloid positivity,^1^ and cut-offs based on three range approach (multinomial regression with *svyVGAM* package)^23^ and tau positivity (binary) as secondary outcomes.^1^ Models with ADPRS included 10 first principal components (PC) of genetic ancestry. Family structure was adjusted for in all models. See eMethods in Supplement 1 for details.

Twin data including MZ and DZ twin pairs can be used to estimate the relative contribution of additive genetic (A) and common (C) and unique (E) environmental effects by decomposing phenotypic variance into these components. The A effects presents a narrow sense heritability (h^2^ = additive genetic variance / phenotypic variance), C effects are all non-genetic effects that make twins within a pair similar, and E effects denote environmental effects that make twins within a pair different including measurement error. We used *OpenMx* structural equation package (2.21.8) in R (eMethods in Supplement 1).^24^

The genome-wide association analyses were performed using mixed linear model using age and sex as covariates with sparse genetic relationship matrix as the random effect of the model controlling for familial and more distant genetic relatedness. The association testing were performed regressing out the covariates from the phenotype and using the adjusted phenotype for the analyses which were performed using genetic complex trait analysis (GCTA)’s^25^ fastGWA function.^26^ After analysis, we filtered out all variants with effect allele frequency < 1%, HWE-p-value < 1e-06 and imputation quality < 0.7. The extended Simes test, GATES,^27^ was used to perform gene-based analyses.

Thresholds for statistical significance were *P* > 5 ^x^ 10^−8^ in the GWAS and P > 5 ^x^ 10^−6^ in the gene-based tests. The regional plot was generated with LocusZoom.^28^

## Results

### Descriptive statistics and associations of age, sex, and education with p-tau217

Mean age (SD) of participants was 76.2 (4.6) years, 57% (n=398) were women, 44% (n=308) were from full twin pairs and 389 were studied without their co-twin. P-tau217 (n=696) ranged from 0.02 to 2.84, the mean (SD) being 0.45 (0.31) pg/mL (Table 1, eFigure1 in Supplement 1). Age was positively associated with p-tau217 (r= 0.24, p <0.001) (Figure 1, eTable1 in Supplement 1). Sex and education were not associated with p-tau217 (Table 1, eTable 2 in Supplement 1).

**Table 1.** Sample demographics.

|  | All | Men | Women | MZ | DZ<sup>c</sup> | Uncertain zygosity |
| --- | --- | --- | --- | --- | --- | --- |
| N (full twin pairs) | 697 (154) | 299 (55) | 398 (99) | 240 (81) | 450 (73) | 7 (0) |
| Age, M (SD), years | 76.17 (4.57) | 76.65 (4.38) | 75.81 (4.68) | 75.24 (5.13) | 76.63 (4.17) | 78.47 (3.50) |
| Education, Median (IQR), years | 10<br>(7–13) | 10<br>(7–18) | 10<br>(7–13) | 10<br>(7–13) | 10<br>(7–13) | 7<br>(7–10) |
| TELE, M (SD), score | 18.87 (1.17) | 18.7 (1.22) | 18.99 (1.12) | 18.79 (1.13) | 18.9<br>(1.2) | 18.93 (0.45) |
| TELE <16<sup>a</sup>, No. (%) | 27 (3.9) | 13 (4.4) | 14 (3.5) | 11 (4.6) | 16 (3.6) | 0 (0) |
| APOE ε4 carriers, No. (%) | 203 (29.2) | 87 (29.2) | 116 (29.1) | 75 (31.4) | 127 (28.2) | 1 (14.3) |
| p-tau217, M (SD), pg/mL | 0.45 (0.31) | 0.47 (0.32) | 0.43 (0.31) | 0.43<br>(0.3) | 0.46 (0.32) | 0.42 (0.40) |
| p-tau217 >0.42<sup>b</sup>, No. (%) | 269 (38.6) | 125 (41.9) | 144 (36.2) | 85 (35.6) | 182 (40.4) | 2 (28.6) |
| p-tau217 <0.40<sup>b</sup>, No. (%) | 407 (58.5) | 162 (54.4) | 245 (61.6) | 144 (60.3) | 258 (57.3) | 5 (71.4) |
| p-tau217 = 0.40 – 0.63<sup>b</sup>, No. (%) | 148 (21.3) | 65 (21.8) | 83 (20.9) | 58 (24.3) | 90 (20) | 0 (0) |
| p-tau217 >0.63<sup>b</sup>, No. (%) | 141 (20.3) | 71 (23.8) | 70 (17.6) | 37 (15.5) | 102 (22.7) | 2 (28.6) |

Abbreviations. TELE = telephone assessment of dementia, APOE = Apolipoprotein E, p-tau217 = phosphorylated tau 217 immunoassay, MZ = monozygotic, DZ = dizygotic.
<sup>a</sup> cut-point of cognitive impairment from Järvenpää et al. 2002. <sup>b</sup> cut-points from Ashton et al. 2024: >0.42 is a threshold for amyloid positivity, <0.40, 0.40 – 0.63, and >0.63 are thresholds for three range approach for classifying individuals into low, intermediate and high probability of amyloid positivity. <sup>c</sup> DZ twins included 313 (53 full pairs) from same-sex twin pairs and 137 (20 full pairs) from opposite-sex twin pairs. One participant had missing data in p-tau217, one participant had missing data in APOE, and two participants had missing data in TELE.

**Figure 1.**
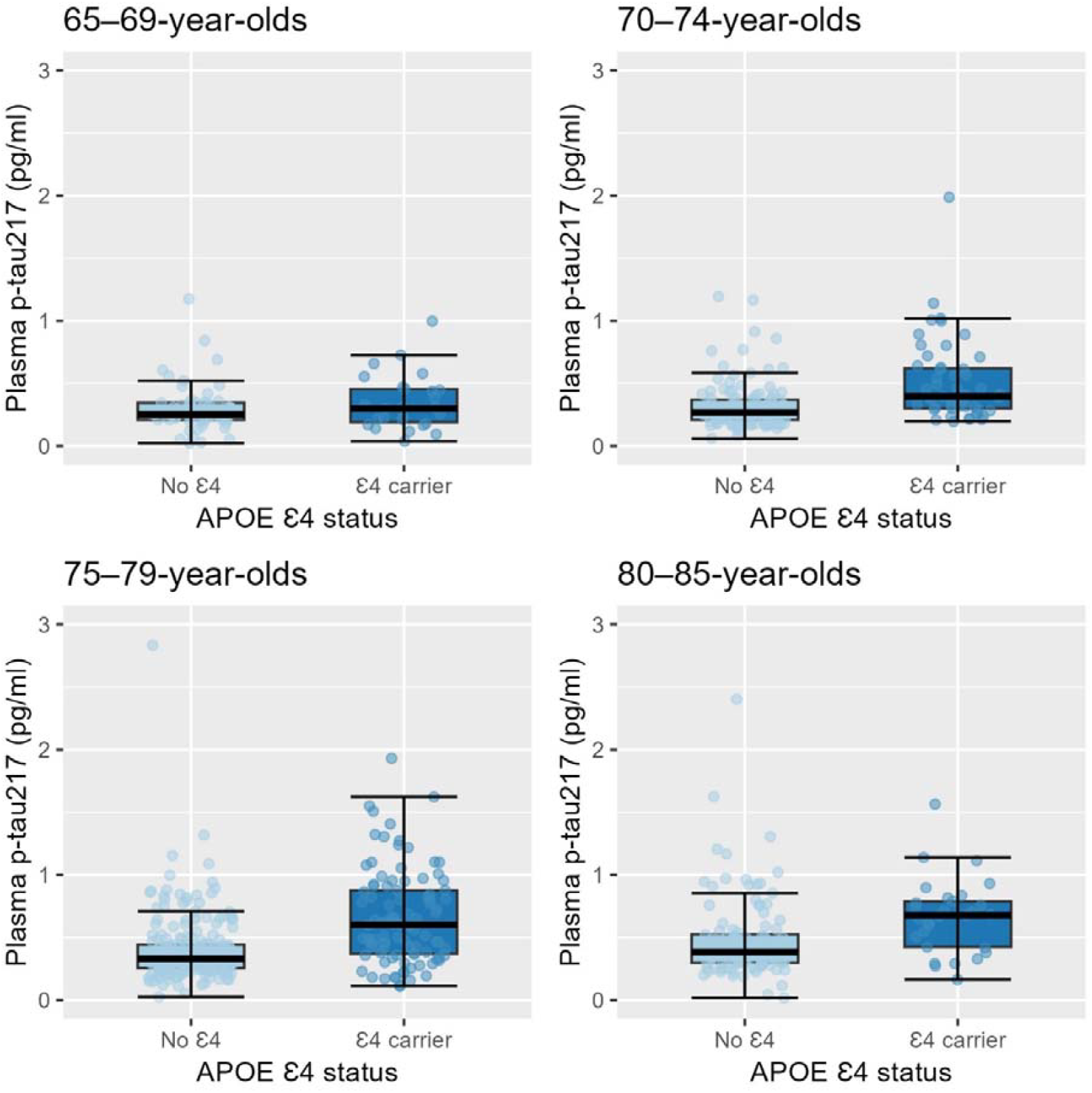
Phosphorylated tau 217 immunoassay levels in *Apolipoprotein* E (*APOE*) ε4-carriers versus non-carriers by age group (n=695).

### ACE models for estimating the heritability and APOE and ADPRS associations with p-tau217

Intra-pair correlations were 0.55 (95%CI, 0.37-0.69) in MZ and 0.24 (95%CI, 0.02-0.46) in DZ pairs. AE model with a heritability estimate of 0.56 (95%CI, 0.36-0.79) was the best fitting model (Table 2). *APOE* ε4-carriers had significantly higher p-tau217 (mean [SD]= 0.58 [0.35] pg/mL) compared to non-carriers (mean [SD]= 0.39 [0.27] pg/mL) (b= 0.41; 95%CI, 0.32-0.51, *P*<0.001, R^2^=0.08) (Figure 1, eTables 2&4 in Supplement 1). ADPRS’s with *APOE* (b=0.03; 95%CI, (−0.02)-0.08, *P* =.193, R^2^=0.001) or without *APOE* (b=0.02; 95%CI, (−0.02-0.07, *P* =.307, R^2^=0.0005) were not significantly related to p-tau217 (eTables 3&4 in Supplement 1).

**Table 2.** Model fitting results for estimating the relative proportion of genetic and environmental effects on phosphorylated tau 217.

| Model | ep | -2LL | df | Δdf | Δ-2LL | P | AIC | A (95% CI) | C (95% CI) | E (95% CI) |
| --- | --- | --- | --- | --- | --- | --- | --- | --- | --- | --- |
| Saturated | 10 | 796.18 | 284 | NA | NA | NA | 816.18 | NA | NA | NA |
| ACE | 4 | 801.98 | 290 | 6 | 5.8 | 0.446 | 809.98 | 0.69 (0.19–1.28) | -0.11 (-0.6–0.3) | 0.43 (0.32–0.61) |
| **AE** | **3** | **802.25** | **291** | **1** | **0.27** | **0.599** | **808.25** | **0.56 (0.36–0.79)** | **NA** | **0.45 (0.33–0.61)** |
| CE | 3 | 809.25 | 291 | 1 | 7.27 | 0.007 | 815.25 | NA | 0.39 (0.23–0.58) | 0.61 (0.49–0.77) |
| E | 2 | 833.33 | 292 | 2 | 31.35 | < 0.001 | 837.33 | NA | NA | 1 (0.85–1.18) |

Abbreviations. ep = number of estimated parameters, -2LL = minus two log likelihood, df = degrees of freedom, $\Delta$ df = change in degrees of freedom, $\Delta$ -2LL = change in minus two log likelihood; AIC = Akaike's information criterion, A = additive genetic effects, C = common environmental effects, E = unique environmental effects. CI = Confidence interval, NA = not applicable. Note that estimate can be negative because we estimated A, C and E components directly from the variances/covariances without any constraints. ACE model was compared against the saturated model. AE, CE, and E models were tested against the ACE model. Models included 77 full monozygotic and 70 full dizygotic twin pairs.

### Prevalence and predictors of AD pathology based on abnormal p-tau217

We used categorical p-tau217 to indicate AD pathology based on previously published cut-offs for beta-amyloid and tau positivity.^1^ The prevalence of AD pathology was 38.6% using >0.42pg/mL cut-off indicative of beta-amyloid positivity and was higher as a function of older age from 20.7% in 65-69 year-olds to 50.7% in 80-85 year-olds (Table 1, eTable 1 in Supplement 1). Older age (OR= 1.15; 95% CI, 1.10-1.20, *P*< 0.001) and having *APOE* ε4-allele (OR=4.53; 95%CI, 3.10-6.62) were predictors of amyloid positivity (eTables 5&6). ADPRS’s with (OR=1.12; 95%CI, 0.95-1.33) or without *APOE* (OR= 1.09; 95%CI, 0.91-1.30 (eTables 5&6 in Supplement 1) were not related to greater odds of amyloid positivity.

The association of *APOE* was even greater when using three range approach (OR=7.22 (95%CI, 4.50-11.59, *P*<0.001) contrasting those with low (p-tau217 <0.40 pg/mL) and high (p-tau217 >0.63 pg/ml) probability of amyloid positivity, and when using >0.64pg/mL binary cut-off indicative of tau positivity (OR=5.13 (95%CI, 3.32-7.91, *P*<0.001) (eTables 7&8 in Supplement 1). The prevalence of tau positivity was 19.6% when using >0.64pg/mL cut-off (eTable 1 in Supplement 1). Sex and education were not associated with abnormal levels of p-tau217 with any cut-offs (eTables 5-8 in Supplement 1).

### Genome-wide association analysis of p-tau217 plasma protein levels

Genomic inflation factor λ was 1.01 indicating no inflation in p-values caused by population stratification or sample relatedness (eFigure 2 in Supplement 1). We found a total of 45 lead variants with genome-wide significant (*P* > 5 ^x^ 10^−8^) associations with p-tau217 blood levels (Figure 2A, Table 3, eTable 9 in Supplement 1), more than a third (17/45) were in chromosome 19 in the *APOE* region (Figure 2B). Top hit was rs429358, a SNP that is used to define *APOE* ε4 carrier status (presence of C-allele, b=0.19, SE=0.02, *P*= 5 ^x^ 10^−16^). In contrast, rs7412 (T-allele indicating presence of AD protective ε2-allele, b= -0.10, SE=0.04, *P*= 0.005) was not among the top SNPs.

**Table 3.**
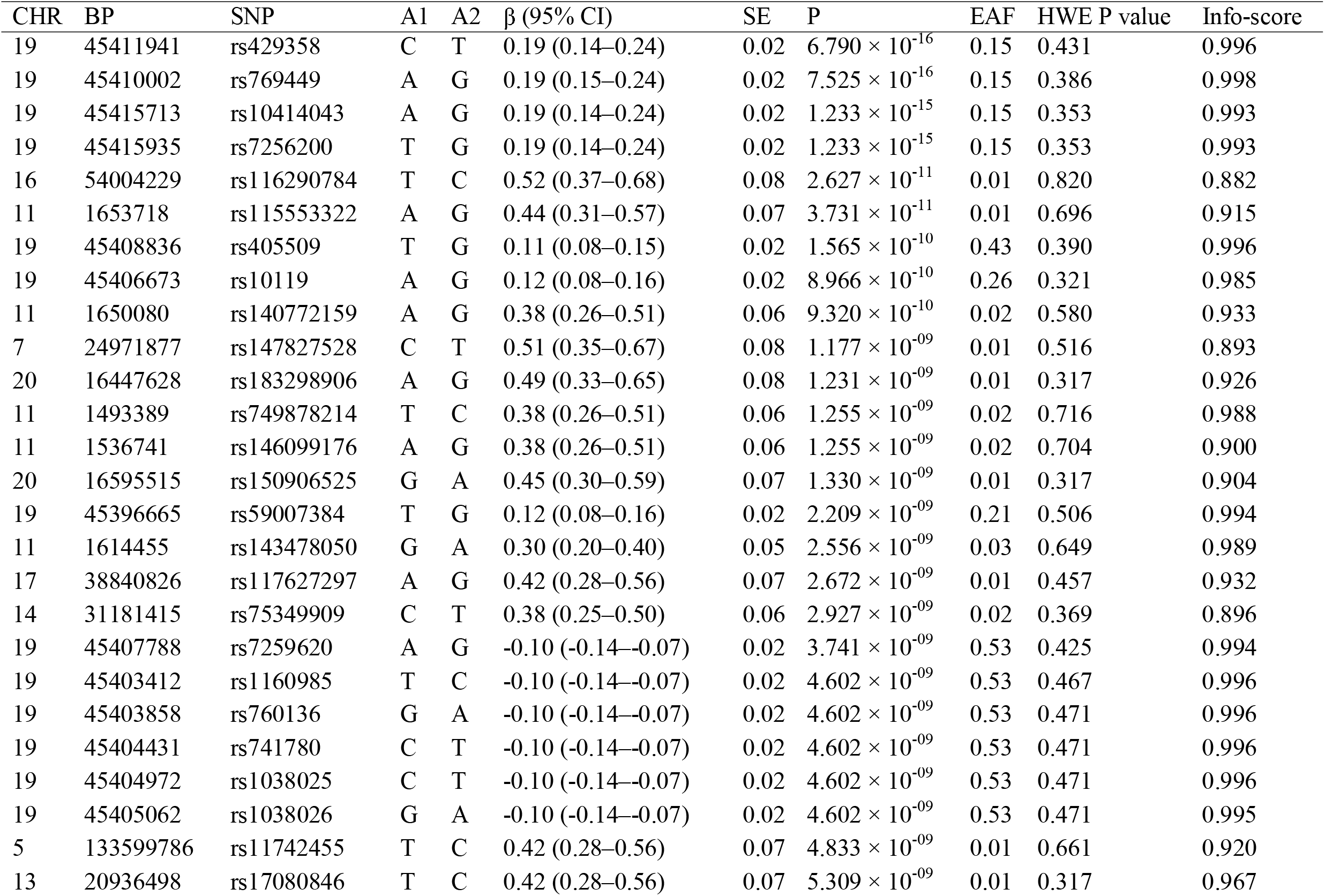
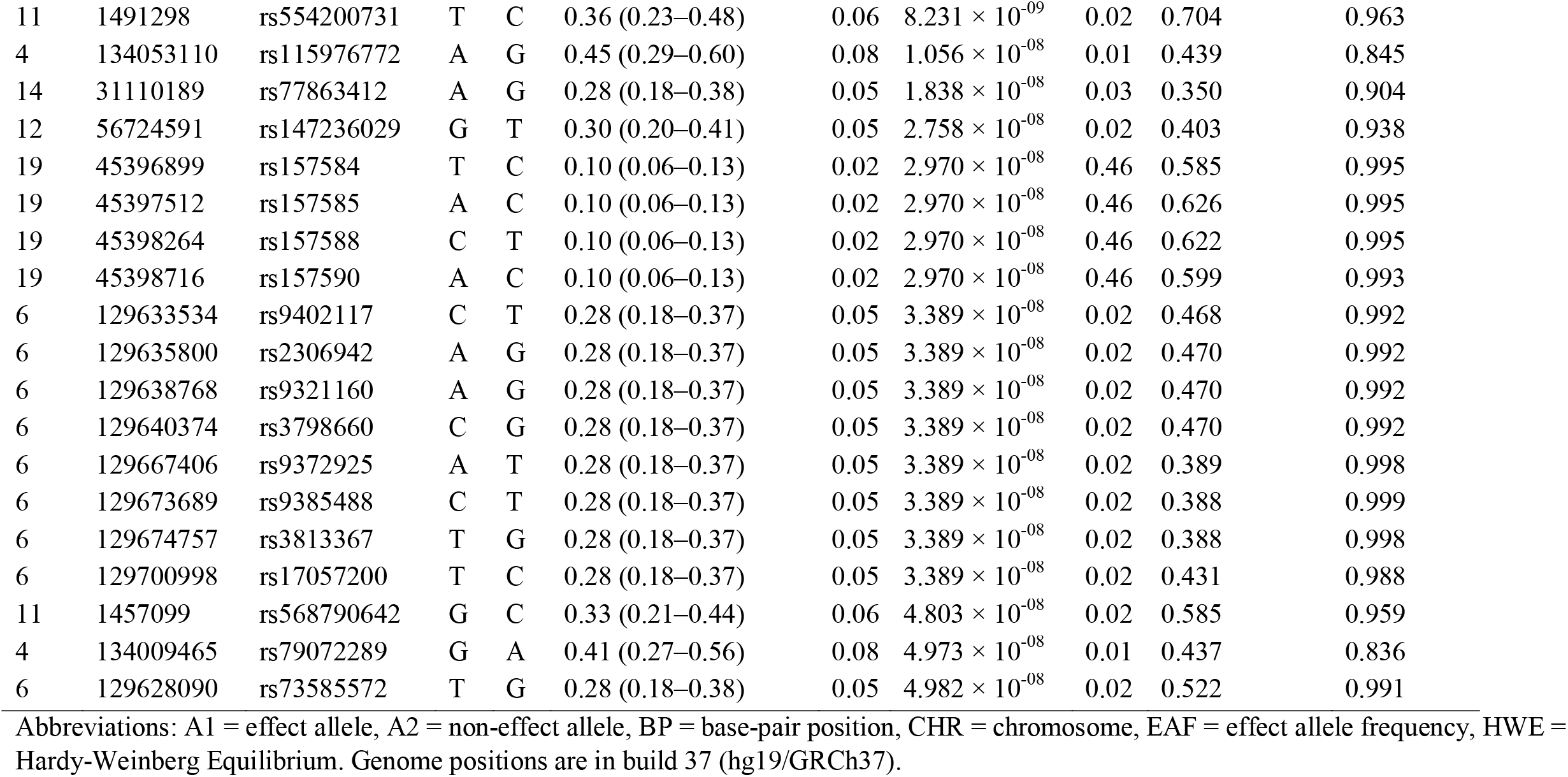
Single nucleotide polymorphisms (SNPs) with P < 5 ^x^ 10^−08^ from the genome-wide association analysis of p-tau217 (n=695).

**Figure 2.**
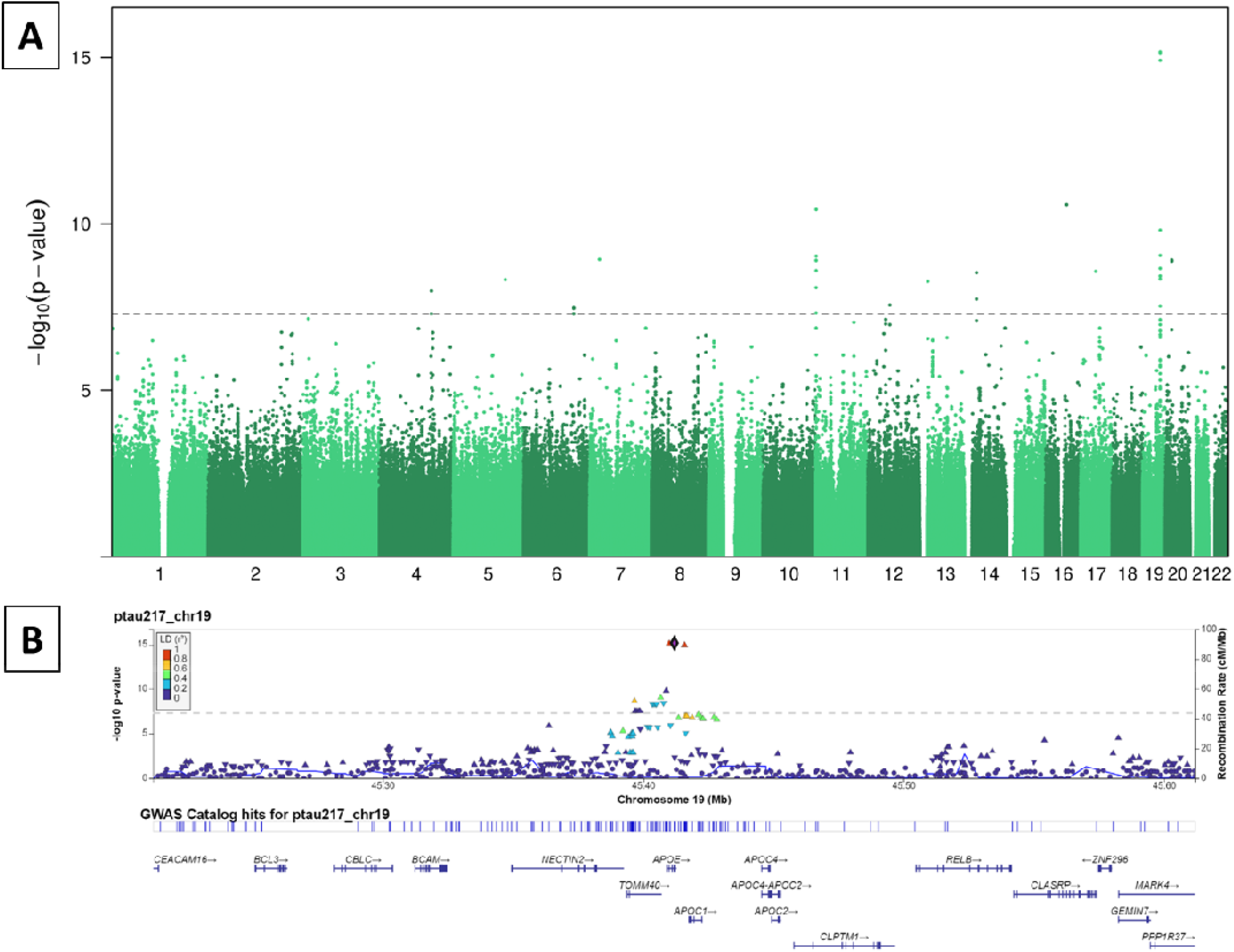
Genome-wide association analysis of p-tau217. A) Manhattan plot for the genome-wide association analysis of p-tau217. B) Regional plot in the apolipoprotein E locus in Chromosome 19 for the single nucleotide polymorphism associations with p-tau217 with rs429358 as a reference.

Gene-based tests indicated five genes (*P* > 5 ^x^ 10^−6^) in the APOE region including *APOE*, translocase of outer mitochondrial membrane 40 *(TOMM40)*, apolipoprotein C1 *(APOC1)*, nectin cell adhesion molecule 2 *(NECTIN2)*, and apolipoprotein C1 pseudogene 1 *(APOC1P1)* (eTable10 in Supplement 1). GCTA based heritability estimate of p-tau217 was 49.2% (SE 43%).

### Post-hoc analyses on APOE genotype subgroups

We investigated the associations of *APOE* risk allele ε4 (ε3/ε4 and ε4/ε4 groups separately), protective ε2 allele (combining four individuals with ε2/ε2 and 52 individuals with ε2/ε3) and the combination of risk and protective alleles ε2/ε4 on p-tau217 (ε3/ε3 as a reference group and age, sex, and education as covariates). Those with one (n=174, b= 0.40[95%CI, 0.30-0.50], *P*<0.001) or two (n=12, b= 0.80[95%CI, 0.49-1.15], *P*<0.001) ε4-alleles had significantly higher p-tau217 than those with two ε3-alleles (n=436). Having ε2-allele was not associated with p-tau217 (n=56, b= -0.11[95%CI, -0.27-0.05], *P*=0.182). Finally, ε2/ε4 group did not differ (n=17, b=0.11 [95%CI, - 0.17-0.40], *P*=0.435) from ε3/ε3 group.

## Discussion

We found that a substantial proportion of the variance in p-tau217 was explained by additive genetic effects with a heritability estimate of 56%. To our knowledge, only one twin study – not including p-tau217 – has investigated the heritability of blood-based AD biomarkers.^4^ We found that age and *APOE* genotype, but not ADPRS (based on cases having clinical diagnosis of AD), were significant predictors of continuous p-tau217 levels but also AD pathology as indicated by abnormal levels of plasma p-tau217 using published cut-offs for amyloid and tau positivity.^1^ For the ADPRS, we chose to use the score based on Lambert et al. (2013).^16^ The larger sample sizes in later genome-wide association studies of AD – all with smaller r-squared than the 0.09 in Lambert et al. – have included also proxy cases where AD status is not based on clinical diagnosis but on family history, thus PRS’s from these studies reflect a more general dementia phenotype that is not specific to AD.^6^

To our knowledge, this was the first genome-wide association study of plasma p-tau217 where we found a total of 45 significant SNP’s. These implicated *APOE* gene but also other chromosome 19 genes in the same region: *TOMM40, APOC1, NECTIN2 and APOC1P1*. Except for *APC1P1*, these genes are protein coding with *APOE* and *APOC1* involved in lipid/cholesterol metabolism, *TOMM40* involved in protein transport into mitochondria and *NECTIN2* as entry for herpesvirus and involved in cell to cell spreading of viruses. *APOE, APOC1* and *TOMM40* are in high linkage disequilibrium, but there is evidence that having additional risk alleles in *APOC1* and *TOMM40* increases the risk of AD over and above the risk of *APOE* ε4-allele.^29^ Together, *APOE, APOC1* and *NECTIN2* haplotypes confirm a risk of AD over and above *APOE*.^30^ Our results suggest broader haplotype in the *APOE* region in association with plasma p-tau217.

The association of *APOE* gene with AD and its cognitive and biomarkers is well-established, but largest GWAS studies using case-control designs including proxy cases have not implicated other *APOE* region genes that were associated with p-tau217 in our study.^31-33^ However, a case-control GWAS study (with family history of dementia as an exclusion criteria in cases) with 9 out of 13 significant SNPs in the *APOE* region implicated *APOE, APOC1, TOMM40* and *NECTIN2* in prediction of clinical diagnosis of AD in Chinese population.^34^ GWAS of p-Tau determined from cerebrospinal fluid samples (CSF) indicated four loci including *APOE*.^35^ *APOE* ε4-allele determining rs429358-C was associated with higher CSF p-Tau levels whereas rs7412-T was associated with lower p-tau. ^35^ In our study, rs429358-C was associated with higher p-tau217 but rs7412-T was not related to p-tau217 levels.

Earlier GWASs of plasma p-tau181 have indicated no significant SNPs other than in the *APOE* gene.^10,11^ In line with our results, GWAS on plasma ApoE found significant SNP’s in the same *APOE* region genes than our study and gene-based tests indicated that *APOE, APOC1, TOMM40* and *NECTIN2* were related to plasma ApoE concentration and risk of AD.^36^ GWAS study of possible preclinical neurodegenerative disease blood biomarkers of caspase-3-cleaved (TAU-C) and ADAM-10 cleaved tau (TAU-A) found only one significant SNP for each of these biomarkers: *APOC1* (rs10414043) for TAU-A and *APOE* (rs429358) for TAU-C. ^37^ These SNPs were our third and first top SNP’s in relation to p-tau217, respectively.

In the biological AT(N)-classification, biomarker classification and cognitive staging are independent and AD is simply defined on biomarker status of amyloid and tau positivity.^7^ Probability for amyloid positivity of 39% in our study was very similar to the prevalence of PET-amyloid positivity in a population-based sample of cognitively healthy persons.^38^ Using biological definition of A+T+, the prevalence of AD pathology in our sample was estimated to be about 20% based on the three range and tau positivity cut-offs. Given our sample excluding individuals with a clinical diagnosis of AD, the substantial proportion of amyloid and tau positive individuals indicate that plasma p-tau217 is useful biomarker for determining AD biological continuum independent of cognitive staging and can guide in screening, diagnosis and prognosis of AD. Together with cognitive staging p-tau217 may help in detecting preclinical (biomarker positive and normal cognition) and prodromal (biomarker positive and mild cognitive impairment) AD.

### Limitations and strengths

Despite our exclusion based on health care registries, about 4% of our sample had cognitive impairment according to telephone screening. Individuals with cognitive impairment may have undiagnosed clinical AD despite not having the diagnosis yet, but we note that scores below the cut-off for cognitive impairment in this telephone-administered instrument may also arise from other reasons than dementia (e.g., hearing problems or environmental disturbances during the interview).

Participation rate of our study was 26%, which can be considered adequate for a clinical biobank recall study requiring participants to travel to the study site for an in-person visit. Strengths included a population-based sample with the expected approximately 30% prevalence of *APOE* ε4-carriership^15^ and health care register-based exclusion of those diagnosed with clinical AD and many other medical conditions affecting the central nervous system. We chose the ADPRS balancing the sample size, proportions of variance explained and criteria of clinically confirmed cases and controls.^6^ To date there is no ADPRS based on biologically defined cases. We did not detect significant associations of ADPRS and *APOE* ε2-allele with p-tau217 (although the direction of association was as expected) which is likely due to relatively sample size. Considering our GWAS, despite multiple significant SNPs associated with p-tau217, our sample was relatively small even for a biomarker study, and we did not have access to a replication sample. However, our results encourage the use of plasma p-tau217 in future GWAS as it is easily scalable measure that captures the biology of AD.

## Conclusions

This cross-sectional population-based study indicated that over half of the variance in p-tau217 was explained by additive genetic effects in individuals without dementia. Age and *APOE*, but not sex were associated with p-tau217 and abnormal levels indicative of AD pathology. Based on previously published cut-offs, many – especially those who were 75-85 years old – had a high probability of AD pathology even though they had no diagnosis of AD. Our GWAS was able to detect polygenic signal for plasma p-tau217 and findings implicated genes in the *APOE* region. Our results support the use of commercially available p-tau217 immunoassay in detecting preclinical or prodromal AD, in large-scale screening of participants for drug trials and lifestyle interventions, and in genome-wide association studies for biologically defined AD.

## Supporting information

FinnGen author list

## Data Availability

TWINGEN data is stored at the THL Biobank for those participants who gave consent for transferring their data to the biobank. Data is available to qualified applicants from academia and companies, for details see https://thl.fi/en/research-and-development/thl-biobank/for-researchers/application-process.

## Acknowledgements

We thank the participants of the older Finnish Twin Cohort study who participated in TWINGEN study. We thank Sabrina Belgasem, Tarja Hallaranta, Henna Palin, Minttu Virolainen, Anna-Kaisa Pohjonen, Anu Outinen-Tuuponen, Marja-Leena Kytökangas, Riikka-Mari Siiro-Virtanen, Senni Lipponen, Nina Hurula, Heidi Kalve, Anniina Friman and biomedical laboratory scientist students from the Turku University of Applied Sciences’ clinical laboratory for collecting the data and processing the blood samples. We thank Finnish Clinical Biobank Tampere, Arctic Biobank, Biobank Borealis of Northern Finland, Central Finland Biobank, Biobank of Eastern Finland and Turku University of Applied Sciences for conducting data collection in Tampere, Oulu, Jyväskylä, Kuopio and Turku, respectively. We thank Auli Toivola and Aija Kyttälä from the THL biobank for study coordination and participant recall, and Anu Jalanko, Huei-Yi Shen, Steffi Besserlink and Jyrki Tammerluoto from the University of Helsinki for study administration and contracting.

## Author contributions

Responsibility for the integrity of the work as a whole, from inception to published article: Vuoksimaa

Concept and design: Runz, Kaprio, Julkunen, Vuoksimaa

Acquisition, analysis or interpretation of data: Saari, Palviainen, Hiltunen, Herukka, Kokkola, Kärkkäinen, Urjansson, Aaltonen, Palotie, Runz, Kaprio, Julkunen, Vuoksimaa

Administrative, technical or material support: Urjansson, Palotie, Kaprio, Julkunen, Vuoksimaa

Drafting of the manuscript: Vuoksimaa

Statistical analysis: Saari, Palviainen

Critical review of the manuscript for important intellectual content: Saari, Palviainen, Hiltunen, Herukka, Kokkola, Kärkkäinen, Urjansson, Aaltonen, Palotie, Runz, Kaprio, Julkunen, Vuoksimaa

Obtained funding: Julkunen, Kaprio, Vuoksimaa

Supervision: Vuoksimaa

