## Supplement 1 for "Cross-sectional study of plasma phosphorylated Tau 217 in persons without dementia"

Contents:

**eMethods**. Pages 2-5

*TWINGEN study design and protocol*

*Measures*

*Ethical approval and informed consents*

*Statistical analyses*

*Structural equation modeling of twin data*

*Genome-wide association study*

**References.** Pages 6-7

**eTables 1-6**. Pages 8-23.

**eTable 1.** P-tau217 values and proportions exceeding cut-off values for amyloid and tau positivity by age groups.

**eTable 2.** Associations of *Apolipoprotein E* (*APOE*) genotype, age, sex and education with p-tau217.

**eTable 3.** Association of Alzheimer’s disease polygenic risk score with p-tau217.

**eTable 4.** Associations of Alzheimer’s disease polygenic risk score and *Apolipoprotein E* genotype with p-tau217.

**eTable 5.** Association of Alzheimer’s disease polygenic risk score with abnormal p-tau217 using >0.42pg/mL cut-off

**eTable 6.** Associations of Alzheimer’s disease polygenic risk score and *Apolipoprotein E* genotype with abnormal p-tau217 using >0.42pg/mL cut-off

**eTable 7.** Predictors of Alzheimer’s disease pathology using three range approach classifying individuals into low, intermediate and high probability of amyloid positivity

**eTable 8.** Predictors of Alzheimer’s disease pathology using p-tau217 >0.64 cut-off pg/mL for tau positivity.

**eTable 9.** Top 100 SNPs (P< 5 ^x^ 10^-08^ are bolded) by Chromosome for genome-wide significant loci associated with plasma pTau-217 (n=695).

**eTable 10.** Top 100 genes (P<5 ^x^ 10^-06^ are bolded) associated with the plasma p-tau217 listed according to p-value (n=695).

**eFigures 1-2.** Pages 24-25.

**eFigure 1**. Distribution of p-tau217 (raw values, residual scores adjusted for age and sex and z-scores of the residual scores).

**eFigure2.** Quantile-quantile plot for the p-tau217 genome-wide association analysis.

**Abbreviation.** p-tau217 = phosphorylated tau 217 immunoassay quantified with ALZpath Simoa pTau-217 v2 Assay Kit (Quanterix, Ref# 104371).

Abnormal p-tau217 >0.42pg/mL and three range approach values for amyloid positivity, and >0.64pg/mL for tau positivity are based on Ashton et al. 2024 cut-offs.^1^

**eMethods**

*TWINGEN study design and protocol*

Finnish Twin Cohort (FTC) study is a longitudinal population-based study of all Finnish same-sex twin pairs born before 1958. The baseline questionnaire was sent in 1975 and multiple follow-up data collections with postal questionnaires have been conducted. In addition to twins from same-sex twin pairs, also twins from opposite-sex twin pairs born in 1938-1949 were sent a baseline questionnaire in 1995-1996. In addition to questionnaire studies, participants of the FTC have been invited to participate in multiple sub-studies also including DNA sample collections for genotyping.^2^ Genotypings in the FTC have been performed in multiple sites including Wellcome Trust Sanger Institute (Cambridge, UK), Institute for Molecular Medicine Finland - FIMM (Helsinki, Finland), University of Chicago Genomics Facility (Chigaco IL, USA), Thermo Fisher Scientific (Santa Clara CA, USA).

In 2018, genome-wide genotyping data and basic characteristics such as body mass-index and smoking status from all FTC participants with available genome-wide genotyping data were transferred to THL biobank according to Finnish biobank law. In the current study, we invited FTC twins born in 1938-1957 and included in the THL biobank to participate in TWINGEN study (see Vuoksimaa, Saari et al. 2023 for details about recruitment).^3^ Invitation letters and consent forms were sent from the THL biobank after checking the eligibility for study participation.

Exclusion was based on health registry data and the exclusion criteria were the following ICD codes: G30 and F00 (Any variant of Alzheimer’s disease or any dementia relating to Alzheimer’s disease, dementia with Lewy bodes, frontotemporal dementia and mixed dementia), F01-F03 (Dementia, any etiology), G20 (Parkinson’s disease), G35 (Multiple sclerosis), I60, I61 and I63 (Intracerebral hemorrhage, subarachnoid hemorrhage or ischemic stroke and their subcategories), S06.1-S06.7 (Traumatic brain injuries other than concussion (S06.0)), F20 (Schizophrenia), F31 (Bipolar disorder), F33, F32.1-F32.3, F34 (Recurrent depression, moderate or severe depression and long-lasting mood disorders), F60 (Personality disorders), F10 (All diagnoses relating to excessive usage of alcohol), F11-F19 (Intoxication because of opioids, cannabinoids, sedative medication, cocaine or hallucinogens), F70–73 (Different stages of intellectual disabilities)

We identified 6053 twins who were born in 1938-1957, who had participated in the population-based older Finnish Twin Cohort (FTC) and who were included in the THL biobank and are also included in the FinnGen study. We note that, additional criterion for participation was inclusion in FinnGen biobank study compiling biobank and health registry data from over 500,000 Finnish citizens who have donated samples to biobank studies in Finland.^4^ After excluding those with AD or other neurodegenerative disease or other cognition affecting disease based on heath registry information, a total of 2718 individuals were invited to participate in TWINGEN study. Number of invited twins was based on the predefined goal of 800 participants returning the informed consent. A total of 830 participants returned signed informed consent for participation and these individuals were contacted by phone to check the suitability and availability for in-person visit to one of the six study sites across Finland. Finally, 697 individuals participated in TWINGEN study yielding a participation rate of 26%. Blood-samples were taken from 696 participants in March 2023 – November 2023. One individual had missing data in genetic analyses.

*Measures*

Education (in years) was based on a self-report questionnaire and chosen from eight categories or a free form answer if none of the eight categories were considered suitable for the participant. If level of education reported in the free form answer corresponded to one of the eight categories, that category was recorded. In cases where a participant chose a level of education from prespecified alternatives and additionally indicated a free-form answer that contradicted their chosen level, the highest level of education was recorded. If the education level remained unclear, highest level of education reported in 1975 or 1981 FTC data collection waves was used; if the level of education was still unclear, the median (10 years of education) was imputed. If the participant had reported higher education in the earlier data collection waves of the FTC study in 1975 or 1981, the highest education level was used. See Silventoinen et al. 2017 for details about educational attainment classes and corresponding years of education.^5^ A dichotomous variable of education years (less than 12 years of education vs. at least 12 years of education) was used in the analyses.

Cognitive status was based on telephone assessment of dementia (TELE) with Finnish cut-off score of <16 for cognitive impairment.^6^ TELE is a cognitive screening instrument that has a correlation of over .8 with the Mini Mental State Examination.^6,7^

Array-based genotyping was used to define *Apolipoprotein E* (*APOE*) genotype status based on imputed single nucleotide polymorphisms of rs429358 and rs7412, a method with 99% accuracy,^8^ and to calculate a polygenic risk score of Alzheimer’s disease (ADPRS).^9^

ADPRS was calculated implementing Bayesian approach to take account the linkage disequilibrium between each SNP utilizing external LD reference panel to adjust the infinitesimal model for polygenic scoring.^10^ The LD reference panel consist of 27284 unrelated samples from the national FINRISK study.^11^ The GWAS summary statistics for Alzheimer’s disease were obtained from The National Institute on Aging Genetics of Alzheimer’s Disease Data Storage Site (https://www.niagads.org/datasets/ng00036) originating from Lambert et al. (2013) study.^9^ There were weights for 54162 samples for the risk score calculation. The LD reference panel, summary statistics and the target study samples of TWINGEN were restricted to HapMap3 (HapMap3 Consortium, 2010) SNPs with European minor allele frequency > 5% and excluding major histocompatibility complex region from chromosome 6. Total number of SNPs used for risk score calculation was 1112327. The genotype data processing, quality control and imputation are described elsewhere.^12^

For the ADPRS, we chose to use the score based on Lambert et al. (2013).^9^ The larger sample sizes in later genome-wide association studies of AD – all with smaller r-squared than the 0.09 in Lambert et al. – have included also proxy cases where AD status is not based on clinical diagnosis but on family history, thus PRS’s from these studies reflect a more general dementia phenotype that is not specific to AD.^13^ We chose the ADPRS balancing the proportion of variance explained, total sample size and the proportion of clinically confirmed cases and controls. To date there is no AD PRS based on biologically defined cases.

Zygosity was based on genotyping data for all participants whose co-twin has also participated and provided DNA sample in earlier data collections of the FTC. If DNA based zygosity was not available we used a questionnaire based measure that has accuracy of over 90%.^14^

*Ethical approval and informed consents*

Ethical approvals for TWINGEN protocol were obtained from the Coordinating Ethics Committee of the Hospital District of Helsinki and Uusimaa (HUS) (number 16831/2022) and THL Biobank approved the research plan with the permission no: THLBB2022_83. All participants gave written informed consent before their participation and had possibility to withdrawn from the study at any point. One consent was given to the University of Helsinki twin study group and the other consent was for THL biobank allowing data transfer from University of Helsinki to THL biobank based on data transfer agreement between the institutions.

*Statistical analyses*

Linear mixed models were used to investigate the associations of age, sex, education *APOE* (ε4 carriers vs. non-carriers and sub-groups in post-hoc analyses) and ADPRS with p-tau217 using the *lme4* package.^15^ R^2^-values for the proportion of variance of p-tau217 explained by *APOE* or ADPRS derived from the univariate models without covariates. We used logistic regression model (*survey* package)^16^ by including all these factors in predicting the probability of amyloid positivity based on validated >0.42pg/ml cut-off.^1^ In addition, we used the *svyVGAM* package^17^ to run a multinomial regression model with three range approach cut-offs to contrast those with low probability (p-tau217 <0.40 pg/mL) of amyloid positivity against the intermediate group (p-tau218 = 0.40-0.63 pg/mL) and high probability group (p-tau217 >0.63 pg/mL).^1^ In addition, we used also p-tau217 >0.64 pg/mL cut-off value for tau positivity.^1^

Models with ADPRS included 10 first principal components (PC) of genetic ancestry. ADPRS and PC’s were standardized for model converge. Family structure was taken into account in all models by including family id as a random effect in linear mixed models and as a cluster variable in logistic and multinomial regression models using *survey* package.^16^ P-tau217 was log was log-transformed in linear mixed models due to its skewness.^18^ Due to missing data for p-tau217 (n=1) and APOE (N=1), the number of participants for models including these variables was 695.

*Structural equation modeling of twin data*

Before running the structural equation models, DZ twins from same- (SSDZ) and opposite-sex (OSDZ) pairs were combined as one group. The difference of within-twin pair correlations of p-tau217 in SSDZ and OSDZ was examined using the *cocor* package.^19^ The within pair correlation of pTau-217 was 0.24 (95% CI: -0.04–0.48) in SSDZ twin pairs and 0.20 (95% CI: -0.28–0.60) in OSDZ twin pairs. The difference in correlations was not statistically significant (z = -0.16, p = 0.871) supporting the use of combined DZ group. Then, we performed a linear regression model with age and sex as predictors of p-tau217 and used the z-transformed residual score in the twin models (excluding those with <-3SD or >+3SD; n=11 participants, of which 4 were MZ, 6 OSDZ and 1 SSDZ individuals from different families yielding 77 MZ and 70 DZ full twin pairs in the models). Before conducting structural equation modeling, we calculated intra-pair correlations of standardized p-tau217 residual score separately for MZ and DZ twin pairs.

We used OpenMx package^20^ version 2.21.8^21^ with NPSOL optimizer^22^ to conduct structural equation modeling for estimating the relative importance of additive genetic (A), and common (C) and unique (E) environmental effects on p-tau217.

First, we ran fully saturated model allowing for different means and variances for MZ and DZ twins and for the twin1 and twin2 (randomly ordered as first and second members of a twin pair). Next, we compared the full ACE model (means and variances constrained to be equal for MZ and DZ and for first and second twins) against the saturated model. After that, we compared reduced AE, CE and E models against the full ACE model to test if constraining of A or C or both, A and C components, to zero resulted in reduced model fit. E effects were included in all models because these include measurement error. Change in model fit was tested by using change in minus 2 log likelihood considering the change in degrees of freedom (using chi-square distribution), whereby p value of less than 0.05 indicated that the component cannot be constrained to zero. Thus p-value ≥0.05 indicated good model fit and the more parsimonious model with fewer estimated parameters was preferred. Model fit was also compared by using Akaike’s information criterion (AIC) values.

Variance components were estimated directly from the variance-covariance matrices without any constraints, thus the estimates can be negative. This approach is less prone to incorrect rejection of A or C effects than the path specification approach where variance components are constrained to have a lower boundary zero by squaring the path coefficients.^22,23^ The direct variance estimation approach also has higher rates of model convergence and smaller bias in parameter estimates.

Finally, using twin data without additional family members, models cannot estimate dominant (D) genetic effects simultaneously with C effects. The initial decision on whether to use ACE or ADE model was based on the comparison of intra-class correlations of MZ and DZ twins. In case of MZ pair correlations being greater than DZ pair correlations, but not greater than double of the DZ correlation, ACE model is the preferred model. If the MZ twin pair correlation is greater than double of the DZ twin pair correlation, then ADE model in indicated.

*Genome-wide association study*

The genome-wide association analyses were performed using mixed linear model using age and sex as covariates with sparse genetic relationship matrix as the random effect of the model controlling for familial and more distant genetic relatedness. The association testing were performed regressing out the covariates from the phenotype and using the adjusted phenotype for the analysis which were performed using genetic complex trait analysis (GCTA)’s^24^ fastGWA function.^25^ After analysis, we filtered out all variants with effect allele frequency < 1%, HWE-pvalue < 1e-06 and imputation quality < 0.7. The extended Simes test, GATES,^26^ were used to perform gene-based analyses.

21.	*_OpenMx: Extended Structural Equation Modelling_. R package version*

*2.21.8*. 2023.

eTable 1. P-tau217 values and proportions exceeding cut-off values for amyloid and tau positivity by age groups (n=696).

| Age group (years) | 65–69 | 70–74 | 75–79 | 80–85 |
| --- | --- | --- | --- | --- |
| n | 82 | 170 | 306 | 138 |
| p-tau217, M (SD), pg/mL | 0.32 (0.20) | 0.38 (0.25) | 0.49 (0.33) | 0.53 (0.36) |
| p-tau217 > 0.42^a^ pg/ml, No. (%) | 17 (20.7) | 42 (24.7) | 140 (45.8) | 70 (50.7) |
| p-tau217 > 0.64^b^ pg/mL, No. (%) | 6 (7.3) | 18 (10.6) | 75 (24.5) | 38 (27.5) |

^a^ cut-off value based on amyloid positivity, and ^b^ cut-off value based on tau positivity from Ashton et al. 2024.^1^

eTable 2. Associations of *Apolipoprotein E* (*APOE*) genotype, age, sex and education with p-tau217 (n=695).

| Predictor | b (95% CI) | *P* |
| --- | --- | --- |
| *APOE* ε4-carrier | 0.41 (0.32–0.51) | < 0.001 |
| Age (centered) | 0.04 (0.03–0.05) | < 0.001 |
| Sex (woman) | -0.06 (-0.15–0.03) | 0.190 |
| Education (≥12 years) | 0.05 (-0.04–0.15) | 0.270 |

Confidence Intervals (CI) and p-values adjusted for clustered family data.

eTable 3. Association of Alzheimer’s disease polygenic risk score (ADPRS) with p-tau217 (n=695).

| Predictor | b (95%CI’s) | *P* |
| --- | --- | --- |
| ADPRS | 0.03 (-0.02–0.08) | 0.193 |
| Age (centered) | 0.04 (0.03–0.05) | < 0.001 |
| Sex (woman) | -0.07 (-0.17–0.02) | 0.122 |
| Education (≥12 years) | 0.03 (-0.07–0.13) | 0.536 |
| PC1 | -0.04 (-0.09–0.01) | 0.090 |
| PC2 | 0.04 (-0.01–0.09) | 0.088 |
| PC3 | 0.01 (-0.03–0.06) | 0.567 |
| PC4 | -0.002 (-0.05-0.05) | 0.915 |
| PC5 | 0.02 (-0.04–0.08) | 0.586 |
| PC6 | 0.02 (-0.04–0.08) | 0.568 |
| PC7 | 0.04 (-0.01–0.09) | 0.121 |
| PC8 | -0.01 (-0.06–0.04) | 0.795 |
| PC9 | -0.02 (-0.08–0.03) | 0.358 |
| PC10 | -0.04 (-0.1–0.01) | 0.147 |

PC = principal component of genetic ancestry. ADPRS and PC’s standardized (M=0, SD=1). Confidence Intervals (CI) and p-values adjusted for clustered family data.

eTable 4. Associations of Alzheimer’s disease polygenic risk score (ADPRS) and *Apolipoprotein E* (*APOE*) genotype with p-tau217 (n=695).

| Predictor | b (95%CI’s) | *P* |
| --- | --- | --- |
| ADPRS without *APOE* | 0.02 (-0.02–0.07) | 0.307 |
| *APOE* ε4-carrier | 0.42 (0.32–0.52) | < 0.001 |
| Age (centered) | 0.04 (0.03–0.05) | < 0.001 |
| Sex (woman) | -0.07 (-0.16–0.02) | 0.130 |
| Education (≥12 years) | 0.04 (-0.05–0.14) | 0.363 |
| PC1 | -0.06 (-0.1– -0.01) | 0.015 |
| PC2 | 0.03 (-0.02–0.07) | 0.200 |
| PC3 | 0.02 (-0.03–0.06) | 0.420 |
| PC4 | -0.01 (-0.05–0.04) | 0.830 |
| PC5 | 0.02 (-0.04–0.08) | 0.499 |
| PC6 | 0.02 (-0.04–0.08) | 0.504 |
| PC7 | 0.04 (-0.01–0.08) | 0.114 |
| PC8 | -0.003 (-0.05–0.05) | 0.897 |
| PC9 | -0.02 (-0.06–0.03) | 0.553 |
| PC10 | -0.03 (-0.08–0.03) | 0.344 |

PC = principal component of genetic ancestry. ADPRS and PC’s standardized (M=0, SD=1). Confidence Intervals (CI) and p-values adjusted for clustered family data.

eTable 5. Association of Alzheimer’s disease polygenic risk score (ADPRS) with abnormal p-tau217 using >0.42pg/mL cut-off (n=695).

| Predictor | OR (95%CI’s) | *P* |
| --- | --- | --- |
| ADPRS | 1.12 (0.95–1.33) | 0.174 |
| Age (centered) | 1.12 (1.08–1.17) | < 0.001 |
| Sex (woman) | 0.81 (0.58–1.13) | 0.218 |
| Education (≥12 years) | 1.06 (0.74–1.52) | 0.764 |
| PC1 | 0.90 (0.76–1.06) | 0.208 |
| PC2 | 1.08 (0.91–1.28) | 0.404 |
| PC3 | 1.11 (0.93–1.32) | 0.237 |
| PC4 | 0.94 (0.78–1.15) | 0.555 |
| PC5 | 1.07 (0.86–1.33) | 0.531 |
| PC6 | 1.01 (0.82–1.26) | 0.895 |
| PC7 | 1.13 (0.95–1.34) | 0.178 |
| PC8 | 1.04 (0.86–1.25) | 0.686 |
| PC9 | 0.90 (0.76–1.08) | 0.266 |
| PC10 | 0.91 (0.78–1.07) | 0.262 |

PC = principal component of genetic ancestry. ADPRS and PC’s standardized (M=0, SD=1). Confidence Intervals (CI) and p-values adjusted for clustered family data.

eTable 6. Associations of Alzheimer’s disease polygenic risk score (ADPRS) and *Apolipoprotein E* (*APOE*) genotype with abnormal p-tau217 using >0.42pg/mL cut-off (n=695).

| Predictor | OR (95% CI) | *P* |
| --- | --- | --- |
| ADPRS without APOE | 1.09 (0.91–1.30) | 0.337 |
| APOE ε4-carrier | 4.53 (3.10–6.62) | < 0.001 |
| Age (centered) | 1.15 (1.10–1.20) | < 0.001 |
| Sex (woman) | 0.80 (0.56–1.14) | 0.216 |
| Education (≥12 years) | 1.14 (0.78–1.67) | 0.503 |
| PC1 | 0.83 (0.70–0.98) | 0.033 |
| PC2 | 1.05 (0.89–1.25) | 0.559 |
| PC3 | 1.15 (0.96–1.36) | 0.124 |
| PC4 | 0.93 (0.76–1.13) | 0.453 |
| PC5 | 1.08 (0.86–1.36) | 0.510 |
| PC6 | 1.02 (0.81–1.28) | 0.887 |
| PC7 | 1.12 (0.94–1.33) | 0.216 |
| PC8 | 1.05 (0.87–1.26) | 0.606 |
| PC9 | 0.93 (0.77–1.12) | 0.445 |
| PC10 | 0.96 (0.81–1.13) | 0.607 |

PC = principal component of genetic ancestry. ADPRS and PC’s standardized (M=0, SD=1). Confidence Intervals (CI) and p-values adjusted for clustered family data.

eTable 7. Predictors of Alzheimer’s disease pathology using three range approach classifying individuals into low, intermediate and high probability of amyloid positivity (n=695).

|  | < 0.40 pg/mL vs 0.40–0.63 pg/mL | | < 0.40 pg/mL vs > 0.63 pg/mL | |
| --- | --- | --- | --- | --- |
| Predictor | OR (95%CI’s) | *P* | OR (95%CI’s) | *P* |
| ADPRS without APOE | 1.08 (0.88–1.33) | 0.468 | 1.16 (0.91–1.47) | 0.229 |
| APOE ε4-carrier | 2.91 (1.84–4.61) | < 0.001 | 7.22 (4.50–11.59) | < 0.001 |
| Age (centered) | 1.11 (1.06–1.17) | < 0.001 | 1.19 (1.12–1.26) | < 0.001 |
| Sex (woman) | 0.86 (0.57–1.31) | 0.487 | 0.64 (0.41–1.00) | 0.050 |
| Education (≥12 years) | 1.26 (0.82–1.94) | 0.298 | 1.09 (0.66–1.77) | 0.744 |
| PC1 | 0.77 (0.62–0.95) | 0.013 | 0.88 (0.70–1.10) | 0.267 |
| PC2 | 1.13 (0.92–1.39) | 0.250 | 1.06 (0.84–1.33) | 0.621 |
| PC3 | 1.03 (0.83–1.27) | 0.777 | 1.18 (0.95–1.47) | 0.136 |
| PC4 | 0.98 (0.78–1.22) | 0.842 | 0.88 (0.70–1.12) | 0.308 |
| PC5 | 0.98 (0.74–1.30) | 0.892 | 1.20 (0.90–1.61) | 0.211 |
| PC6 | 0.98 (0.73–1.33) | 0.910 | 1.12 (0.85–1.49) | 0.414 |
| PC7 | 1.05 (0.86–1.29) | 0.631 | 1.22 (0.96–1.54) | 0.102 |
| PC8 | 0.94 (0.74–1.18) | 0.570 | 1.03 (0.82–1.30) | 0.792 |
| PC9 | 0.93 (0.73–1.19) | 0.582 | 0.96 (0.76–1.21) | 0.727 |
| PC10 | 0.98 (0.79–1.22) | 0.858 | 0.87 (0.68–1.11) | 0.259 |

ADPRS = Alzheimer's disease polygenic risk score, APOE = Apolipoprotein E, PC = principal component of genetic ancestry. ADPRS and PC’s have been standardized (M=0, SD=1). Confidence Intervals (CI) and p-values adjusted for clustered family data. The group with p-tau217 < 0.40 pg/mL (low probability) was used as the reference in contrasts against 0.40-0.63pg/mL = intermediate probability, and >0.63pg/mL = high probability of amyloid positivity.

eTable 8. Predictors of Alzheimer’s disease pathology using p-tau217 >0.64 cut-off pg/mL for tau positivity (n=695).

| Predictor | OR (95%CI’s) | *P* |
| --- | --- | --- |
| ADPRS without APOE | 1.13 (0.89–1.44) | 0.303 |
| APOE ε4-carrier | 5.13 (3.32–7.91) | < 0.001 |
| Age (centered) | 1.15 (1.09–1.22) | < 0.001 |
| Sex (woman) | 0.74 (0.48–1.13) | 0.158 |
| Education (≥12 years) | 0.96 (0.59–1.54) | 0.857 |
| PC1 | 0.94 (0.76–1.18) | 0.611 |
| PC2 | 1.01 (0.81–1.26) | 0.919 |
| PC3 | 1.14 (0.93–1.40) | 0.220 |
| PC4 | 0.90 (0.72–1.13) | 0.352 |
| PC5 | 1.22 (0.92–1.61) | 0.164 |
| PC6 | 1.12 (0.85–1.48) | 0.405 |
| PC7 | 1.21 (0.96–1.53) | 0.112 |
| PC8 | 1.07 (0.85–1.34) | 0.567 |
| PC9 | 0.95 (0.76–1.18) | 0.653 |
| PC10 | 0.86 (0.66–1.12) | 0.256 |

Abbreviations. ADPRS = Alzheimer's disease polygenic risk score, APOE = Apolipoprotein E, PC = principal component of genetic ancestry. Confidence Intervals (CI) and p-values adjusted for clustered family data. ADPRS and PC’s have been standardized (M=0, SD=1). Number of individuals with p-tau217 >0.64pg/mL was 137 (19.7%).

**eTable 9.** Top 100 SNPs (P< 5 ^x^ 10^-08^ are bolded) by Chromosome for genome-wide significant loci associated with plasma pTau-217 (n=695).

| CHR | BP | SNP | A1 | A2 | β (95% CI) | SE | P | EAF | HWE P value | Info-score |
| --- | --- | --- | --- | --- | --- | --- | --- | --- | --- | --- |
| 1 | 3196053 | rs1574225 | T | C | 0.43 (0.27–0.59) | 0.08 | 1.397 × 10^-07^ | 0.01 | 0.809 | 0.981 |
| 2 | 188728879 | rs190774966 | G | T | 0.42 (0.26–0.58) | 0.08 | 1.835 × 10^-07^ | 0.01 | 0.493 | 0.978 |
| 2 | 216510492 | rs75567970 | T | C | 0.33 (0.20–0.45) | 0.06 | 1.988 × 10^-07^ | 0.02 | 0.420 | 0.927 |
| 2 | 213556085 | rs113890773 | C | G | 0.41 (0.25–0.56) | 0.08 | 2.187 × 10^-07^ | 0.01 | 0.485 | 0.927 |
| 3 | 14679590 | rs74674539 | A | G | 0.35 (0.23–0.48) | 0.07 | 7.025 × 10^-08^ | 0.02 | 0.969 | 0.911 |
| **4** | **134053110** | **rs115976772** | **A** | **G** | **0.45 (0.29–0.60)** | **0.08** | **1.056 × 10^-08^** | **0.01** | **0.439** | **0.845** |
| **4** | **134009465** | **rs79072289** | **G** | **A** | **0.41 (0.27–0.56)** | **0.08** | **4.973 × 10^-08^** | **0.01** | **0.437** | **0.836** |
| 4 | 100769194 | rs528273556 | T | C | 0.39 (0.24–0.53) | 0.07 | 1.414 × 10^-07^ | 0.01 | 0.708 | 0.936 |
| 4 | 137577895 | rs114238523 | G | C | 0.27 (0.17–0.37) | 0.05 | 1.861 × 10^-07^ | 0.02 | 0.382 | 0.936 |
| 4 | 137599897 | rs75181895 | G | A | 0.27 (0.17–0.37) | 0.05 | 1.861 × 10^-07^ | 0.02 | 0.381 | 0.938 |
| **5** | **133599786** | **rs11742455** | **T** | **C** | **0.42 (0.28–0.56)** | **0.07** | **4.833 × 10^-09^** | **0.01** | **0.661** | **0.920** |
| **6** | **129633534** | **rs9402117** | **C** | **T** | **0.28 (0.18–0.37)** | **0.05** | **3.389 × 10^-08^** | **0.02** | **0.468** | **0.992** |
| **6** | **129635800** | **rs2306942** | **A** | **G** | **0.28 (0.18–0.37)** | **0.05** | **3.389 × 10^-08^** | **0.02** | **0.470** | **0.992** |
| **6** | **129638768** | **rs9321160** | **A** | **G** | **0.28 (0.18–0.37)** | **0.05** | **3.389 × 10^-08^** | **0.02** | **0.470** | **0.992** |
| **6** | **129640374** | **rs3798660** | **C** | **G** | **0.28 (0.18–0.37)** | **0.05** | **3.389 × 10^-08^** | **0.02** | **0.470** | **0.992** |
| **6** | **129667406** | **rs9372925** | **A** | **T** | **0.28 (0.18–0.37)** | **0.05** | **3.389 × 10^-08^** | **0.02** | **0.389** | **0.998** |
| **6** | **129673689** | **rs9385488** | **C** | **T** | **0.28 (0.18–0.37)** | **0.05** | **3.389 × 10^-08^** | **0.02** | **0.388** | **0.999** |
| **6** | **129674757** | **rs3813367** | **T** | **G** | **0.28 (0.18–0.37)** | **0.05** | **3.389 × 10^-08^** | **0.02** | **0.388** | **0.998** |
| **6** | **129700998** | **rs17057200** | **T** | **C** | **0.28 (0.18–0.37)** | **0.05** | **3.389 × 10^-08^** | **0.02** | **0.431** | **0.988** |
| **6** | **129628090** | **rs73585572** | **T** | **G** | **0.28 (0.18–0.38)** | **0.05** | **4.982 × 10^-08^** | **0.02** | **0.522** | **0.991** |
| **7** | **24971877** | **rs147827528** | **C** | **T** | **0.51 (0.35–0.67)** | **0.08** | **1.177 × 10^-09^** | **0.01** | **0.516** | **0.893** |
| 7 | 144358244 | rs17170428 | C | T | 0.30 (0.19–0.41) | 0.06 | 1.343 × 10^-07^ | 0.02 | 0.453 | 0.958 |
| 8 | 140373573 | rs78369189 | A | G | 0.42 (0.26–0.58) | 0.08 | 2.237 × 10^-07^ | 0.01 | 0.551 | 0.974 |
| 8 | 140373709 | rs146194401 | C | T | 0.42 (0.26–0.58) | 0.08 | 2.237 × 10^-07^ | 0.01 | 0.551 | 0.975 |
| 8 | 140376067 | rs143548348 | C | T | 0.42 (0.26–0.58) | 0.08 | 2.237 × 10^-07^ | 0.01 | 0.556 | 0.970 |
| 8 | 140379932 | rs146301213 | A | G | 0.42 (0.26–0.58) | 0.08 | 2.237 × 10^-07^ | 0.01 | 0.556 | 0.980 |
| 8 | 140380318 | rs148087909 | G | A | 0.42 (0.26–0.58) | 0.08 | 2.237 × 10^-07^ | 0.01 | 0.552 | 0.978 |
| 8 | 140382232 | rs145438081 | A | G | 0.42 (0.26–0.58) | 0.08 | 2.237 × 10^-07^ | 0.01 | 0.551 | 0.973 |
| 8 | 140384341 | rs139109722 | G | A | 0.42 (0.26–0.58) | 0.08 | 2.237 × 10^-07^ | 0.01 | 0.557 | 0.979 |
| 8 | 140387548 | rs73723223 | C | T | 0.42 (0.26–0.58) | 0.08 | 2.237 × 10^-07^ | 0.01 | 0.552 | 0.980 |
| 8 | 140387560 | rs73723224 | C | T | 0.42 (0.26–0.58) | 0.08 | 2.237 × 10^-07^ | 0.01 | 0.552 | 0.980 |
| 8 | 140388086 | rs113718680 | T | G | 0.42 (0.26–0.58) | 0.08 | 2.237 × 10^-07^ | 0.01 | 0.552 | 0.981 |
| 8 | 140388127 | rs74958239 | C | T | 0.42 (0.26–0.58) | 0.08 | 2.237 × 10^-07^ | 0.01 | 0.552 | 0.981 |
| 8 | 140388266 | rs74814227 | A | C | 0.42 (0.26–0.58) | 0.08 | 2.237 × 10^-07^ | 0.01 | 0.551 | 0.981 |
| 8 | 140388513 | rs113419225 | A | G | 0.42 (0.26–0.58) | 0.08 | 2.237 × 10^-07^ | 0.01 | 0.551 | 0.981 |
| 8 | 140389022 | rs113162766 | T | C | 0.42 (0.26–0.58) | 0.08 | 2.237 × 10^-07^ | 0.01 | 0.551 | 0.981 |
| 8 | 140389166 | rs74873123 | C | G | 0.42 (0.26–0.58) | 0.08 | 2.237 × 10^-07^ | 0.01 | 0.551 | 0.981 |
| 8 | 140389363 | rs78361115 | A | C | 0.42 (0.26–0.58) | 0.08 | 2.237 × 10^-07^ | 0.01 | 0.551 | 0.981 |
| 8 | 140389480 | rs112242142 | T | C | 0.42 (0.26–0.58) | 0.08 | 2.237 × 10^-07^ | 0.01 | 0.551 | 0.981 |
| 8 | 140389635 | rs111978143 | T | G | 0.42 (0.26–0.58) | 0.08 | 2.237 × 10^-07^ | 0.01 | 0.551 | 0.982 |
| 8 | 140389832 | rs111352692 | G | A | 0.42 (0.26–0.58) | 0.08 | 2.237 × 10^-07^ | 0.01 | 0.551 | 0.981 |
| 8 | 140390507 | rs76237047 | A | G | 0.42 (0.26–0.58) | 0.08 | 2.237 × 10^-07^ | 0.01 | 0.551 | 0.981 |
| 8 | 140390741 | rs112841303 | G | C | 0.42 (0.26–0.58) | 0.08 | 2.237 × 10^-07^ | 0.01 | 0.551 | 0.981 |
| **11** | **1653718** | **rs115553322** | **A** | **G** | **0.44 (0.31–0.57)** | **0.07** | **3.731 × 10^-11^** | **0.01** | **0.696** | **0.915** |
| **11** | **1650080** | **rs140772159** | **A** | **G** | **0.38 (0.26–0.51)** | **0.06** | **9.320 × 10^-10^** | **0.02** | **0.580** | **0.933** |
| **11** | **1493389** | **rs749878214** | **T** | **C** | **0.38 (0.26–0.51)** | **0.06** | **1.255 × 10^-09^** | **0.02** | **0.716** | **0.988** |
| **11** | **1536741** | **rs146099176** | **A** | **G** | **0.38 (0.26–0.51)** | **0.06** | **1.255 × 10^-09^** | **0.02** | **0.704** | **0.900** |
| **11** | **1614455** | **rs143478050** | **G** | **A** | **0.30 (0.20–0.40)** | **0.05** | **2.556 × 10^-09^** | **0.03** | **0.649** | **0.989** |
| **11** | **1491298** | **rs554200731** | **T** | **C** | **0.36 (0.23–0.48)** | **0.06** | **8.231 × 10^-09^** | **0.02** | **0.704** | **0.963** |
| **11** | **1457099** | **rs568790642** | **G** | **C** | **0.33 (0.21–0.44)** | **0.06** | **4.803 × 10^-08^** | **0.02** | **0.585** | **0.959** |
| 11 | 98848591 | rs74885179 | T | A | 0.35 (0.22–0.48) | 0.07 | 9.095 × 10^-08^ | 0.02 | 0.530 | 0.969 |
| 11 | 98877140 | rs144789261 | T | A | 0.35 (0.22–0.48) | 0.07 | 9.095 × 10^-08^ | 0.02 | 0.773 | 0.978 |
| 11 | 1582533 | rs141825788 | G | A | 0.24 (0.15–0.33) | 0.05 | 1.353 × 10^-07^ | 0.03 | 0.803 | 0.996 |
| **12** | **56724591** | **rs147236029** | **G** | **T** | **0.30 (0.20–0.41)** | **0.05** | **2.758 × 10^-08^** | **0.02** | **0.403** | **0.938** |
| 12 | 45913313 | rs74595894 | C | T | 0.27 (0.17–0.37) | 0.05 | 7.354 × 10^-08^ | 0.03 | 0.345 | 0.958 |
| 12 | 45898723 | rs116890891 | A | G | 0.27 (0.17–0.37) | 0.05 | 1.030 × 10^-07^ | 0.03 | 0.345 | 0.965 |
| 12 | 56643342 | rs7296038 | T | C | 0.29 (0.18–0.40) | 0.05 | 1.075 × 10^-07^ | 0.02 | 0.375 | 0.899 |
| 12 | 41085303 | rs11177826 | T | G | 0.41 (0.25–0.56) | 0.08 | 1.990 × 10^-07^ | 0.01 | 0.466 | 0.950 |
| 12 | 41088664 | rs11177844 | A | G | 0.41 (0.25–0.56) | 0.08 | 1.990 × 10^-07^ | 0.01 | 0.466 | 0.949 |
| 12 | 41118245 | rs11178069 | G | A | 0.41 (0.25–0.56) | 0.08 | 1.990 × 10^-07^ | 0.01 | 0.469 | 0.892 |
| **13** | **20936498** | **rs17080846** | **T** | **C** | **0.42 (0.28–0.56)** | **0.07** | **5.309 × 10^-09^** | **0.01** | **0.317** | **0.967** |
| **14** | **31181415** | **rs75349909** | **C** | **T** | **0.38 (0.25–0.50)** | **0.06** | **2.927 × 10^-09^** | **0.02** | **0.369** | **0.896** |
| **14** | **31110189** | **rs77863412** | **A** | **G** | **0.28 (0.18–0.38)** | **0.05** | **1.838 × 10^-08^** | **0.03** | **0.350** | **0.904** |
| 14 | 31054340 | rs77810288 | G | A | 0.26 (0.17–0.36) | 0.05 | 8.142 × 10^-08^ | 0.03 | 0.386 | 0.915 |
| 14 | 31074198 | rs139554593 | A | G | 0.26 (0.17–0.36) | 0.05 | 8.142 × 10^-08^ | 0.03 | 0.386 | 0.918 |
| 14 | 31088207 | rs45573134 | G | C | 0.26 (0.17–0.36) | 0.05 | 8.142 × 10^-08^ | 0.03 | 0.386 | 0.913 |
| 14 | 104708270 | rs139920092 | A | G | 0.43 (0.27–0.59) | 0.08 | 1.347 × 10^-07^ | 0.01 | 0.877 | 0.891 |
| **16** | **54004229** | **rs116290784** | **T** | **C** | **0.52 (0.37–0.68)** | **0.08** | **2.627 × 10^-11^** | **0.01** | **0.820** | **0.882** |
| **17** | **38840826** | **rs117627297** | **A** | **G** | **0.42 (0.28–0.56)** | **0.07** | **2.672 × 10^-09^** | **0.01** | **0.457** | **0.932** |
| 17 | 49309802 | rs573309406 | C | G | 0.40 (0.25–0.55) | 0.08 | 1.338 × 10^-07^ | 0.01 | 0.440 | 0.964 |
| **19** | **45411941** | **rs429358** | **C** | **T** | **0.19 (0.14–0.24)** | **0.02** | **6.790 × 10^-16^** | **0.15** | **0.431** | **0.996** |
| **19** | **45410002** | **rs769449** | **A** | **G** | **0.19 (0.15–0.24)** | **0.02** | **7.525 × 10^-16^** | **0.15** | **0.386** | **0.998** |
| **19** | **45415713** | **rs10414043** | **A** | **G** | **0.19 (0.14–0.24)** | **0.02** | **1.233 × 10^-15^** | **0.15** | **0.353** | **0.993** |
| **19** | **45415935** | **rs7256200** | **T** | **G** | **0.19 (0.14–0.24)** | **0.02** | **1.233 × 10^-15^** | **0.15** | **0.353** | **0.993** |
| **19** | **45408836** | **rs405509** | **T** | **G** | **0.11 (0.08–0.15)** | **0.02** | **1.565 × 10^-10^** | **0.43** | **0.390** | **0.996** |
| **19** | **45406673** | **rs10119** | **A** | **G** | **0.12 (0.08–0.16)** | **0.02** | **8.966 × 10^-10^** | **0.26** | **0.321** | **0.985** |
| **19** | **45396665** | **rs59007384** | **T** | **G** | **0.12 (0.08–0.16)** | **0.02** | **2.209 × 10^-09^** | **0.21** | **0.506** | **0.994** |
| **19** | **45407788** | **rs7259620** | **A** | **G** | **-0.10 (-0.14–-0.07)** | **0.02** | **3.741 × 10^-09^** | **0.53** | **0.425** | **0.994** |
| **19** | **45403412** | **rs1160985** | **T** | **C** | **-0.10 (-0.14–-0.07)** | **0.02** | **4.602 × 10^-09^** | **0.53** | **0.467** | **0.996** |
| **19** | **45403858** | **rs760136** | **G** | **A** | **-0.10 (-0.14–-0.07)** | **0.02** | **4.602 × 10^-09^** | **0.53** | **0.471** | **0.996** |
| **19** | **45404431** | **rs741780** | **C** | **T** | **-0.10 (-0.14–-0.07)** | **0.02** | **4.602 × 10^-09^** | **0.53** | **0.471** | **0.996** |
| **19** | **45404972** | **rs1038025** | **C** | **T** | **-0.10 (-0.14–-0.07)** | **0.02** | **4.602 × 10^-09^** | **0.53** | **0.471** | **0.996** |
| **19** | **45405062** | **rs1038026** | **G** | **A** | **-0.10 (-0.14–-0.07)** | **0.02** | **4.602 × 10^-09^** | **0.53** | **0.471** | **0.995** |
| **19** | **45396899** | **rs157584** | **T** | **C** | **0.10 (0.06–0.13)** | **0.02** | **2.970 × 10^-08^** | **0.46** | **0.585** | **0.995** |
| **19** | **45397512** | **rs157585** | **A** | **C** | **0.10 (0.06–0.13)** | **0.02** | **2.970 × 10^-08^** | **0.46** | **0.626** | **0.995** |
| **19** | **45398264** | **rs157588** | **C** | **T** | **0.10 (0.06–0.13)** | **0.02** | **2.970 × 10^-08^** | **0.46** | **0.622** | **0.995** |
| **19** | **45398716** | **rs157590** | **A** | **C** | **0.10 (0.06–0.13)** | **0.02** | **2.970 × 10^-08^** | **0.46** | **0.599** | **0.993** |
| 19 | 45421254 | rs12721046 | A | G | 0.11 (0.07–0.15) | 0.02 | 7.532 × 10^-08^ | 0.23 | 0.395 | 0.977 |
| 19 | 45416178 | rs483082 | T | G | 0.11 (0.07–0.15) | 0.02 | 1.071 × 10^-07^ | 0.21 | 0.372 | 0.992 |
| 19 | 45416741 | rs438811 | T | C | 0.11 (0.07–0.15) | 0.02 | 1.071 × 10^-07^ | 0.21 | 0.358 | 0.991 |
| 19 | 45422160 | rs12721051 | G | C | 0.10 (0.06–0.14) | 0.02 | 1.536 × 10^-07^ | 0.24 | 0.354 | 0.977 |
| 19 | 45427125 | rs111789331 | A | T | 0.10 (0.07–0.14) | 0.02 | 1.553 × 10^-07^ | 0.23 | 0.361 | 0.970 |
| 19 | 45418790 | rs5117 | C | T | 0.11 (0.07–0.14) | 0.02 | 1.621 × 10^-07^ | 0.21 | 0.373 | 0.987 |
| 19 | 45413576 | rs75627662 | T | C | 0.11 (0.07–0.15) | 0.02 | 1.678 × 10^-07^ | 0.20 | 0.317 | 0.995 |
| 19 | 45422846 | rs56131196 | A | G | 0.10 (0.06–0.14) | 0.02 | 2.121 × 10^-07^ | 0.24 | 0.405 | 0.976 |
| 19 | 45422946 | rs4420638 | G | A | 0.10 (0.06–0.14) | 0.02 | 2.121 × 10^-07^ | 0.24 | 0.405 | 0.976 |
| **20** | **16447628** | **rs183298906** | **A** | **G** | **0.49 (0.33–0.65)** | **0.08** | **1.231 × 10^-09^** | **0.01** | **0.317** | **0.926** |
| **20** | **16595515** | **rs150906525** | **G** | **A** | **0.45 (0.30–0.59)** | **0.07** | **1.330 × 10^-09^** | **0.01** | **0.317** | **0.904** |
| 20 | 16379756 | rs4813219 | T | C | 0.36 (0.23–0.50) | 0.07 | 1.523 × 10^-07^ | 0.02 | 0.679 | 0.938 |
| 20 | 16410060 | rs761902 | G | A | 0.36 (0.23–0.50) | 0.07 | 1.523 × 10^-07^ | 0.02 | 0.562 | 0.951 |

A1 = effect allele. A2 = non-effect allele. BP = base-pair position. CHR = chromosome. EAF = effect allele frequency. HWE = Hardy-Weinberg Equilibrium. SNP = single nucleotide polymorphism.

**eTable 10.** Top 100 genes (P<5 ^x^ 10^-06^ are bolded) associated with the plasma p-tau217 listed according to p-value (n=695).

| Gene | number of SNPs used for *P*-value estimation | GATES *P*-value | Chromosome | Position | Smallest SNP  *P*-value in the region |
| --- | --- | --- | --- | --- | --- |
| ***APOE*** | **17** | **6.787 × 10^-15^** | **19** | **45411941** | **6.790 × 10^-16^** |
| ***TOMM40*** | **22** | **7.965 × 10^-15^** | **19** | **45411941** | **6.790 × 10^-16^** |
| ***APOC1*** | **14** | **1.153 × 10^-14^** | **19** | **45415713** | **1.233 × 10^-15^** |
| ***NECTIN2*** | **68** | **8.870 × 10^-08^** | **19** | **45396665** | **2.209 × 10^-09^** |
| ***APOC1P1*** | **15** | **1.643 × 10^-06^** | **19** | **45427125** | **1.553 × 10^-07^** |
| *OSER1-DT* | 28 | 4.687 × 10^-05^ | 20 | 42844854 | 2.485 × 10^-06^ |
| *NCOA4* | 10 | 5.075 × 10^-05^ | 10 | 51571131 | 6.581 × 10^-06^ |
| *LOC100288846* | 6 | 5.667 × 10^-05^ | 14 | 39733360 | 1.105 × 10^-05^ |
| *TIMM23* | 6 | 6.463 × 10^-05^ | 10 | 51587237 | 1.291 × 10^-05^ |
| *ARID1B* | 224 | 1.231 × 10^-04^ | 6 | 157420255 | 8.811 × 10^-07^ |
| *TIMM23B* | 26 | 1.821 × 10^-04^ | 10 | 51571131 | 6.581 × 10^-06^ |
| *TIMM23B-AGAP6* | 29 | 1.821 × 10^-04^ | 10 | 51571131 | 6.581 × 10^-06^ |
| *NFIB* | 627 | 1.864 × 10^-04^ | 9 | 14457483 | 3.742 × 10^-07^ |
| *ZNF296* | 8 | 1.958 × 10^-04^ | 19 | 45582402 | 2.996 × 10^-05^ |
| *MIMT1* | 10 | 2.358 × 10^-04^ | 19 | 57350608 | 3.289 × 10^-05^ |
| *FITM2* | 10 | 2.441 × 10^-04^ | 20 | 42933577 | 3.079 × 10^-05^ |
| *PEG3-AS1* | 8 | 2.497 × 10^-04^ | 19 | 57323016 | 3.836 × 10^-05^ |
| *NSD1* | 22 | 2.765 × 10^-04^ | 5 | 176581739 | 1.887 × 10^-05^ |
| *LINC02238* | 36 | 2.860 × 10^-04^ | 1 | 74208548 | 1.239 × 10^-05^ |
| *IFITM10* | 15 | 3.000 × 10^-04^ | 11 | 1751657 | 2.917 × 10^-05^ |
| *UBTFL1* | 1 | 3.018 × 10^-04^ | 11 | 89822001 | 3.018 × 10^-04^ |
| *LINC01164* | 22 | 3.452 × 10^-04^ | 10 | 133603551 | 2.831 × 10^-05^ |
| *PEG3* | 14 | 3.810 × 10^-04^ | 19 | 57323016 | 3.836 × 10^-05^ |
| *GEMIN7* | 20 | 3.828 × 10^-04^ | 19 | 45582402 | 2.996 × 10^-05^ |
| *NCAN* | 22 | 4.126 × 10^-04^ | 19 | 19336608 | 2.895 × 10^-05^ |
| *MIA2* | 71 | 4.161 × 10^-04^ | 14 | 39845921 | 1.313 × 10^-05^ |
| *TTC41P* | 59 | 4.529 × 10^-04^ | 12 | 104248696 | 1.638 × 10^-05^ |
| *GDAP1L1* | 16 | 4.565 × 10^-04^ | 20 | 42878144 | 4.229 × 10^-05^ |
| *LINC01019* | 78 | 5.120 × 10^-04^ | 5 | 3472585 | 1.190 × 10^-05^ |
| *CUL2* | 13 | 5.431 × 10^-04^ | 10 | 35339061 | 5.788 × 10^-05^ |
| *BBIP1* | 10 | 5.720 × 10^-04^ | 10 | 112673555 | 7.732 × 10^-05^ |
| *LINC01480* | 5 | 6.181 × 10^-04^ | 19 | 42045576 | 1.754 × 10^-04^ |
| *ZIM2* | 27 | 6.215 × 10^-04^ | 19 | 57323016 | 3.836 × 10^-05^ |
| *COLEC10* | 101 | 6.412 × 10^-04^ | 8 | 119997745 | 1.242 × 10^-05^ |
| *APOA4* | 9 | 6.559 × 10^-04^ | 11 | 116694055 | 9.983 × 10^-05^ |
| *ALDH1A2* | 64 | 6.702 × 10^-04^ | 15 | 58246998 | 3.175 × 10^-05^ |
| *MKRN3* | 6 | 6.832 × 10^-04^ | 15 | 23817721 | 1.787 × 10^-04^ |
| *PDILT* | 41 | 6.965 × 10^-04^ | 16 | 20414902 | 2.593 × 10^-05^ |
| *PEAR1* | 26 | 7.008 × 10^-04^ | 1 | 156869047 | 5.675 × 10^-05^ |
| *LINC00836* | 78 | 7.810 × 10^-04^ | 10 | 26012605 | 1.450 × 10^-05^ |
| *C4A* | 6 | 7.907 × 10^-04^ | 6 | 31944851 | 1.628 × 10^-04^ |
| *LOC110384692* | 6 | 7.907 × 10^-04^ | 6 | 31944851 | 1.628 × 10^-04^ |
| *ERICH4* | 12 | 8.277 × 10^-04^ | 19 | 41950297 | 9.171 × 10^-05^ |
| *WNT9A* | 33 | 8.357 × 10^-04^ | 1 | 228112465 | 3.879 × 10^-05^ |
| *ZDHHC19* | 56 | 8.547 × 10^-04^ | 3 | 195919984 | 2.416 × 10^-05^ |
| *MIR569* | 5 | 8.572 × 10^-04^ | 3 | 170824128 | 2.695 × 10^-04^ |
| *BPIFA2* | 4 | 9.083 × 10^-04^ | 20 | 31746183 | 2.323 × 10^-04^ |
| *DMAC2* | 13 | 9.183 × 10^-04^ | 19 | 41950297 | 9.171 × 10^-05^ |
| *PCLAF* | 9 | 9.329 × 10^-04^ | 15 | 64679594 | 1.578 × 10^-04^ |
| *ATP2B2* | 398 | 9.426 × 10^-04^ | 3 | 10409897 | 5.376 × 10^-06^ |
| *FAM99A* | 32 | 9.477 × 10^-04^ | 11 | 1690911 | 4.169 × 10^-05^ |
| *ATP6V1A* | 6 | 9.815 × 10^-04^ | 3 | 113462123 | 2.140 × 10^-04^ |
| *NAA50* | 6 | 1.068 × 10^-03^ | 3 | 113430599 | 2.183 × 10^-04^ |
| *SPOP* | 24 | 1.076 × 10^-03^ | 17 | 47701415 | 6.818 × 10^-05^ |
| *TRIP4* | 8 | 1.087 × 10^-03^ | 15 | 64679594 | 1.578 × 10^-04^ |
| *SLC29A2* | 4 | 1.103 × 10^-03^ | 11 | 66126169 | 3.139 × 10^-04^ |
| *RDH5* | 6 | 1.104 × 10^-03^ | 12 | 56115785 | 2.265 × 10^-04^ |
| *GOLGA6L7* | 4 | 1.124 × 10^-03^ | 15 | 29096698 | 3.643 × 10^-04^ |
| *DSCR10* | 7 | 1.175 × 10^-03^ | 21 | 39574717 | 2.481 × 10^-04^ |
| *DXO* | 9 | 1.177 × 10^-03^ | 6 | 31944851 | 1.628 × 10^-04^ |
| *TNXB* | 40 | 1.216 × 10^-03^ | 6 | 32017242 | 6.462 × 10^-05^ |
| *GALNT15* | 98 | 1.326 × 10^-03^ | 3 | 16213469 | 2.335 × 10^-05^ |
| *CD63* | 7 | 1.338 × 10^-03^ | 12 | 56115785 | 2.265 × 10^-04^ |
| *C4B* | 13 | 1.399 × 10^-03^ | 6 | 31944851 | 1.628 × 10^-04^ |
| *C4B_2* | 13 | 1.399 × 10^-03^ | 6 | 31944851 | 1.628 × 10^-04^ |
| *NFIA* | 230 | 1.441 × 10^-03^ | 1 | 61786398 | 1.049 × 10^-05^ |
| *NUDT2* | 12 | 1.449 × 10^-03^ | 9 | 34342881 | 1.595 × 10^-04^ |
| *STK19* | 11 | 1.459 × 10^-03^ | 6 | 31944851 | 1.628 × 10^-04^ |
| *ZNF480* | 28 | 1.517 × 10^-03^ | 19 | 52801386 | 8.722 × 10^-05^ |
| *MARCHF1* | 508 | 1.562 × 10^-03^ | 4 | 164538841 | 1.009 × 10^-05^ |
| *SNORD18B* | 9 | 1.572 × 10^-03^ | 15 | 66790490 | 2.067 × 10^-04^ |
| *SNORD16* | 9 | 1.572 × 10^-03^ | 15 | 66790490 | 2.067 × 10^-04^ |
| *DDX11-AS1* | 36 | 1.574 × 10^-03^ | 12 | 31171776 | 8.738 × 10^-05^ |
| *GLRA1* | 34 | 1.582 × 10^-03^ | 5 | 151252935 | 8.030 × 10^-05^ |
| *CCL27* | 10 | 1.585 × 10^-03^ | 9 | 34666093 | 2.662 × 10^-04^ |
| *MSX2P1* | 10 | 1.586 × 10^-03^ | 17 | 56237602 | 2.716 × 10^-04^ |
| *CNPY2* | 21 | 1.587 × 10^-03^ | 12 | 56703722 | 2.147 × 10^-04^ |
| *NOD1* | 36 | 1.634 × 10^-03^ | 7 | 30470423 | 7.843 × 10^-05^ |
| *BLOC1S1* | 9 | 1.642 × 10^-03^ | 12 | 56115785 | 2.265 × 10^-04^ |
| *BLOC1S1-RDH5* | 9 | 1.642 × 10^-03^ | 12 | 56115785 | 2.265 × 10^-04^ |
| *MIR5008* | 6 | 1.655 × 10^-03^ | 1 | 228124414 | 3.762 × 10^-04^ |
| *LOC730098* | 11 | 1.695 × 10^-03^ | 9 | 34666093 | 2.662 × 10^-04^ |
| *ALMS1P1* | 27 | 1.696 × 10^-03^ | 2 | 73875541 | 1.017 × 10^-04^ |
| *OR4D1* | 13 | 1.828 × 10^-03^ | 17 | 56237602 | 2.716 × 10^-04^ |
| *OSER1* | 26 | 1.844 × 10^-03^ | 20 | 42840537 | 1.412 × 10^-04^ |
| *IL17RE* | 17 | 1.859 × 10^-03^ | 3 | 9941613 | 1.899 × 10^-04^ |
| *NT5DC3* | 67 | 1.865 × 10^-03^ | 12 | 104239445 | 4.932 × 10^-05^ |
| *NINJ2-AS1* | 31 | 1.954 × 10^-03^ | 12 | 736859 | 9.912 × 10^-05^ |
| *MIR4508* | 6 | 1.973 × 10^-03^ | 15 | 23807942 | 4.057 × 10^-04^ |
| *MYOM1* | 186 | 1.985 × 10^-03^ | 18 | 3067599 | 1.804 × 10^-05^ |
| *GAB4* | 55 | 1.995 × 10^-03^ | 22 | 17446157 | 5.938 × 10^-05^ |
| *LINC00693* | 180 | 1.997 × 10^-03^ | 3 | 28742974 | 4.468 × 10^-05^ |
| *DCC* | 643 | 2.010 × 10^-03^ | 18 | 49863549 | 1.639 × 10^-05^ |
| *DSCR8* | 10 | 2.052 × 10^-03^ | 21 | 39494241 | 2.774 × 10^-04^ |
| *LOC100134391* | 53 | 2.071 × 10^-03^ | 17 | 71732131 | 6.812 × 10^-05^ |
| *SNORA70I* | 9 | 2.098 × 10^-03^ | 2 | 215710269 | 3.165 × 10^-04^ |
| *MIR6083* | 9 | 2.109 × 10^-03^ | 3 | 124096880 | 3.149 × 10^-04^ |
| *TTLL12* | 54 | 2.138 × 10^-03^ | 22 | 43573834 | 6.056 × 10^-05^ |
| *IL11RA* | 14 | 2.149 × 10^-03^ | 9 | 34666093 | 2.662 × 10^-04^ |
| *LINC02655* | 15 | 2.170 × 10^-03^ | 10 | 82410987 | 2.171 × 10^-04^ |

SNP = single nucleotide polymorphism.

**eFigure1.** Distribution of p-tau217 (raw values, residual scores adjusted for age and sex and z-scores of the residual scores) (n=696).

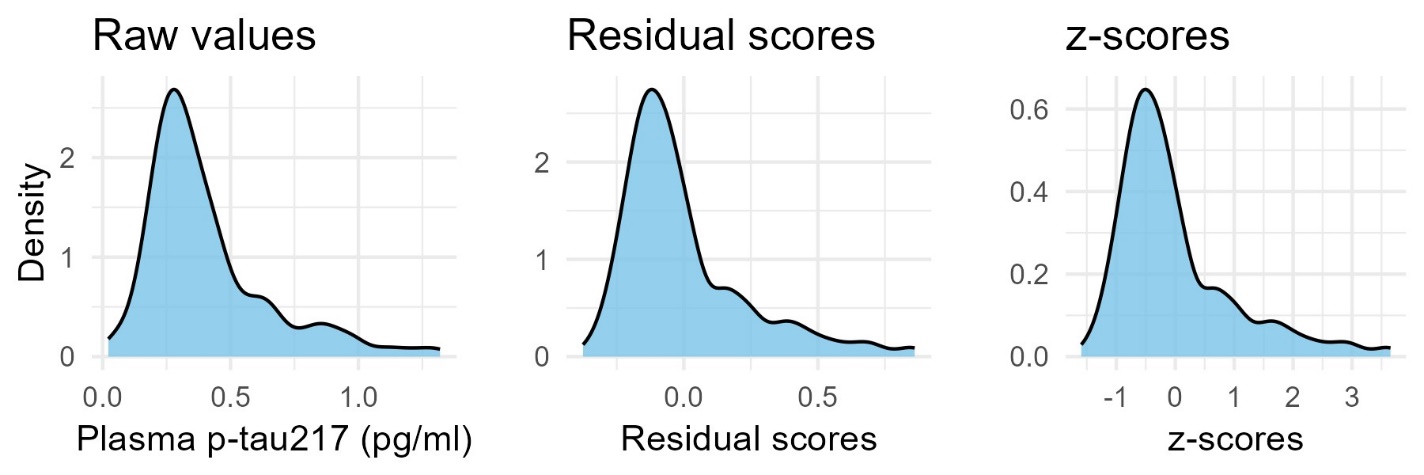

**eFigure 2.** Quantile-quantile plot for the p-tau217 genome-wide association analysis.

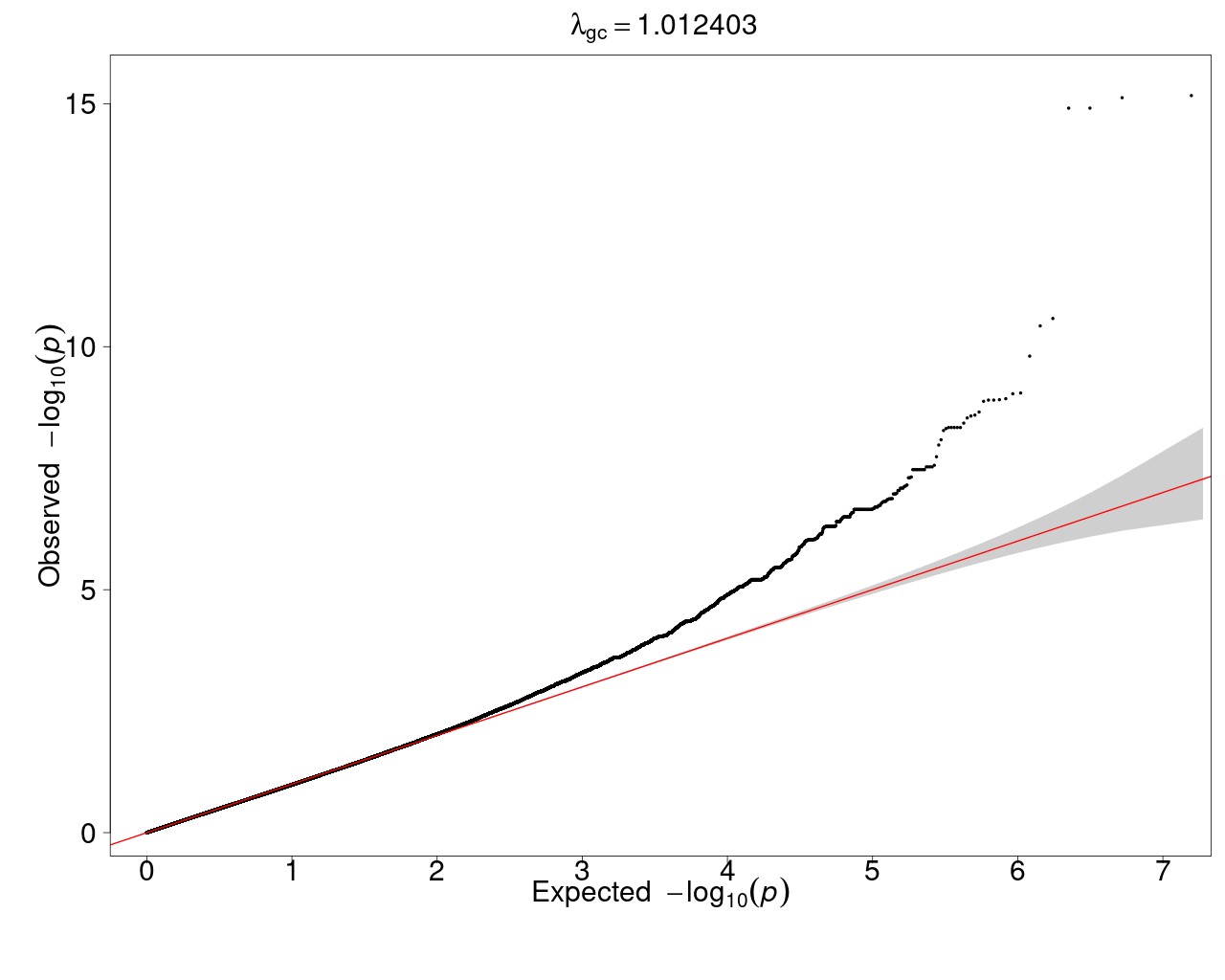
